# Characterizing amyloid and tau PET-based stages across the clinical continuum

**DOI:** 10.1101/2025.09.26.25336751

**Authors:** Karly A. Cody, Andrzej Sokołowski, Emily Johns, Lucah Medina Guerra, Joseph R. Winer, Christina B. Young, Kyan Younes, Logan Dumitrescu, Derek B. Archer, Alaina Durant, Aditi Sathe, Mary Ellen I. Koran, Jesse Mez, Andrew J. Saykin, Arthur Toga, Michael Cuccaro, Duygu Tosun, Philip S. Insel, Sterling C. Johnson, Theresa M. Harrison, Timothy J. Hohman, Elizabeth C. Mormino, A4 and ADNI Study Teams

**Affiliations:** Department of Neurology and Neurological Sciences, Stanford University, Stanford, CA, 94305, USA; Vanderbilt Memory and Alzheimer’s Center, Vanderbilt University Medical Center, Nashville, TN, 37203, USA; Department of Radiology and Radiological Sciences, Vanderbilt University Medical Center, Nashville, TN, 37203, USA; Department of Radiology, Mayo Clinic of Arizona, Scottsdale, AZ, 85259, USA; Department of Neurology, Boston University Chobanian & Avedisian School of Medicine, Boston, MA, 02118, USA; Indiana Alzheimer’s Disease Research Center, Department of Radiology and Imaging Sciences and Department of Medical and Molecular Genetics, Indiana University School of Medicine, Indianapolis, IN, 46202, USA; Alzheimer’s Disease Research Center, Keck School of Medicine, University of Southern California, Los Angeles, CA, 90033, USA; Department of Human Genetics, University of Miami, Miami, FL, 33146, USA; Department of Radiology and Biomedical Imaging, University of California San Francisco, San Francisco, CA, 94143, USA; Department of Psychiatry and Behavioral Sciences, University of California, San Francisco, CA, 94143, USA; Alzheimer’s Disease Research Center, Department of Medicine, University of Wisconsin, Madison, WI, 53705, USA; Neuroscience Department, University of California-Berkeley, Berkeley, CA, 94720, USA; Wu Tsai Neuroscience Institute, Stanford University, Stanford, CA, 94305, USA; Molecular Imaging Program at Stanford, Stanford University, Stanford, CA, 94305, USA

**Keywords:** Amyloid, Tau, PET, Alzheimer’s disease

## Abstract

Staging the severity of Alzheimer’s disease pathology using biomarkers is central to early detection and therapeutic trial design. In this cross-sectional study, we standardized amyloid and tau PET data across multiple cohorts to characterize the frequency of amyloid and tau PET-based stages across the clinical continuum.

We examined amyloid and tau severity in 10,396 participants (mean [SD] age, 71.9 [7.1] years) with amyloid PET imaging and a subset (n = 3,295) with tau PET imaging. Clinical stage was defined using cohort-specific criteria and categorized as cognitively unimpaired (n = 7,764), mild cognitive impairment (n = 1,480), or dementia (n = 1,152). Amyloid positivity was defined as ≥25 centiloids and amyloid severity was staged using centiloids bins (e.g., <10, 10-24, 25-49, 50-74, 75-99, ≥100). Tau PET severity was staged using a hierarchical Braak staging schema (e.g., T−, T12+, T34+, T56+), and combined with amyloid status to operationalize PET-based Alzheimer’s disease biological stages (e.g., Stage A: A+T−; Stage B: A+T12+; Stage C: A+T34+; Stage D: A+T56+). The cumulative probabilities of PET-based stages were estimated using ordinal logistic regression models.

In cognitively unimpaired individuals, the frequency of amyloid levels ≥10 centiloids increased with age. Similarly, amyloid levels ≥25 centiloids increased with age in mild cognitive impairment. Overall, elevated amyloid (≥25 CL) was more likely with increasing age among non-demented individuals. By contrast, this age association was attenuated in dementia where severe amyloid burden (e.g., ≥100 CL) was most common. In the tau PET subsample (n = 3,295), there was a three-way interaction between amyloid, age, and clinical impairment on likelihood of tau severity. Both higher amyloid and greater clinical impairment were associated with increased tau severity; however, the strength and direction of these associations varied with age. At lower amyloid levels, the odds of tau severity increased with older age among cognitively unimpaired and mild cognitive impairment. Conversely, at higher amyloid levels, the odds of higher tau severity (e.g., T56+) decreased with older age in mild cognitive impairment and dementia. A similar age-related pattern was observed in the frequency of biological stages (n = 1,154), where Stage D (e.g., A+T56+) was most frequent in younger individuals with dementia.

These findings underscore the dual importance of amyloid and tau PET severity as biomarkers for staging and characterizing Alzheimer’s disease progression. They also demonstrate the feasibility of applying PET-based staging frameworks for the diagnosis of Alzheimer’s disease across multiple tracers and cohorts.

## Introduction

The pathophysiological processes of Alzheimer’s disease — β-amyloid plaques and tau neurofibrillary tangles (NFTs) — begin decades before symptoms of clinical impairment are present. Advances in PET imaging have demonstrated that abnormal amyloid typically becomes evident first, followed by the accumulation and spread of tau outside the medial temporal lobe which is associated with clinical progression and cognitive impairment.^1–4^ Although amyloid and tau PET are often classified dichotomously as “negative” or “positive”, the continuum of amyloid and tau PET burden have been shown to predict clinical and pathological disease progression.^3,5–10^ Thus, staging the severity of amyloid and tau PET levels has several important implications for early detection and clinical prognosis.

The Centiloid (CL) scale, a standardized measure of global brain amyloid PET levels, ranges from ∼0 CL corresponding to the mean amyloid PET burden of a healthy young adult to ∼100 CL corresponding to the mean amyloid PET burden of patients with mild Alzheimer’s disease dementia.^11^ In research, amyloid levels of <10 CL have been associated with a lack of amyloid pathology at autopsy, ∼25 CL has been shown to agree with visual assessment, and ∼50 CL and higher are often associated with brain tau deposition and clinical disease progression.^9,12–14^ In clinical trial settings, CLs have been used to inform patient selection and stratification as well as dosing modifications with recent trials using CLs to target intervention at early and intermediate levels of amyloid.^15,16^

Similarly, tau PET severity is increasingly used to define disease stage and inform therapeutic strategies. For example, the recent TRAILBLAZER trial enrolled individuals with intermediate tau PET levels, based on the hypothesis that anti-amyloid therapies may be less effective when brain tau burden is too severe.^17^ Tau PET accumulation generally follows a hierarchical spatial pattern of accumulation (e.g. Braak staging)^18–21^, progressing from the medial temporal lobe in early stages to widespread neocortical involvement in later stages. Correspondingly, the Alzheimer’s Association recently proposed a revised framework for staging Alzheimer’s disease severity^22^ which defines four biological stages reflecting progressively greater spatial spread of tau among amyloid positive (A+) individuals. As anti-amyloid therapies become increasingly available in clinical practice, and additional therapeutic pathways^23^ continue to be actively investigated, it is essential to characterize both brain amyloid and tau severity to improve disease staging, identify therapeutic windows, and evaluate therapeutic efficacy.

Staging amyloid and tau severity is also important for clinical prognosis. Individuals with greater than ∼50 CLs have a significantly higher risk of progression to mild cognitive impairment (MCI) or dementia.^9,14^ Higher amyloid and higher tau PET burden is associated with faster preclinical cognitive decline^6,21,24–27^ as well as clinical disease progression.^26,28–30^ However, there remains considerable heterogeneity in the level of brain tau given amyloid,^21,31^ and the level of impairment given the level of amyloid and tau pathology.^21,32,33^ As amyloid and tau PET are increasingly being integrated into therapeutic trial design and clinical care,^17,34,35^ characterizing this heterogeneity will be crucial for understanding and staging Alzheimer’s disease progression.

The widespread integration of PET imaging into aging and Alzheimer’s disease research cohorts over the last decade provides a unique opportunity to explore the extent of amyloid and tau burden and operationalize the revised biological staging criteria using large-scale PET data. In this study, we applied standardized image processing pipelines and thresholding methods across four cohorts to stage amyloid and tau PET severity. We additionally combined these measures to operationalize PET-based Alzheimer’s disease biological stages consistent with the recently proposed biological staging framework.^22^ Finally, we characterized the frequency of amyloid and tau PET-based stages in individuals spanning the clinical continuum.

## Materials and methods

### Participants

Cohorts included the Anti-Amyloid Treatment in Asymptomatic Alzheimer’s disease (A4; https://atri.usc.edu/study/a4-study/), the Alzheimer’s disease Neuroimaging Initiative (ADNI; https://adni.loni.usc.edu/), Alzheimer’s disease Research Centers (ADRCs) in collaboration with the National Alzheimer’s Coordinating Center (NACC; https://naccdata.org/), and the Wisconsin Registry for Alzheimer’s Prevention (WRAP; https://wrap.wisc.edu/).^36^ Participant clinical stage was determined using cohort-specific diagnoses (Supplementary Methods) and included cognitively unimpaired (CU; n = 7,764), mild cognitive impairment (MCI; n = 1,480), and dementia (n = 1,152). Participants with concurrent (+/− 3 years) PET and clinical visits were included in the current study (Supplementary Fig. 1). Written informed consent was obtained from all participants, and local institutional review boards for human research approved the study. The study was performed in accordance with the ethical standards described in the 1964 Declaration of Helsinki and its later amendments or comparable ethical standards.

### PET Image Processing and Quantification

Brain amyloid was assessed using 18F-florbetapir, 18F-florbetaben, or 11C-Pittsburgh Compound B PET imaging. A subset of individuals with amyloid PET imaging also underwent tau PET imaging using 18F-Flortaucipir or 18F-MK-6240 within 3 years of their amyloid scan (Supplementary Fig. 1). Prior to processing, scan acquisition timing underwent quality control, such that only scans with data within 10 minutes before or after the designated recommended acquisition start time for each radiotracer were included (Supplemental Table 1; Supplementary Fig. 1). All scans with standardized acquisition timing underwent MRI-free processing locally at Stanford, using methods consistent with the SCAN MRI-free processing pipelines described by Landau et al.^37^ Broadly, this process included: (1) linearly aligning summed late-frame PET data to an MNI152 T1 template, (2) performing non-linear spatial adjustments to align the summed PET data to amyloid or tau PET template(s), (3) extracting signal intensities from cortical regions of interest (ROIs), and (4) generating standardized uptake value ratios (SUVRs). For amyloid PET, a “universal” amyloid PET template was used for spatial normalization and SUVRs were generated using the GAAIN atlas, which includes a large cortical target region (frontal, parietal, cingulate, and lateral temporal regions) and is commonly used to calculate global centiloids (CLs).^11^ Global cortical SUVRs were then translated to CLs using tracer-specific equations (Supplemental Table 2). For tau PET, multiple tau templates were integrated into the non-linear spatial normalization step to reduce errors in spatial normalization since tau binding patterns are more focal than amyloid and off-target binding varies across different ligands (Supplementary Fig. 2).^38^ Bilateral tau PET SUVRs (gray matter inferior cerebellum reference region) were extracted in composite ROIs approximating Braak neurofibrillary tangle staging^39,40^ defined on a normalized atlas^41^ (Supplementary Fig. 3). Finally, post-processing quality control was conducting using a combination of statistical and visual inspection approaches (Supplemental Methods).

### Amyloid and tau PET positivity and staging

Global amyloid PET positivity (A+/−) was determined as ≥25 CL. Amyloid severity was staged using 6 CL-based bins based on previously published^9,12,13,15,42–46^ studies: <10 CL (negative), 10–24 CL (low/equivocal), 25–49 CL (moderate), 50–74 CL(moderate-high), 75-100 CL (high), ≥100 CL (very high; Figure 1A). Regional tau positivity (T+/−) across Braak-based ROIs was determined using a tracer-specific gaussian mixture model approach (Supplementary Methods; Supplementary Figs. 4-8, Supplementary Table 3). Regional tau PET Z-scores were calculated using the mean and standard deviation of the tracer-specific gaussian mixture model-derived T− distribution. Hierarchical tau PET stages were determined based on sequential positivity across Braak-based ROIs, where later stages could only be achieved if the individual was tau positive in previous stages.^19,21^ Cases that were tau negative across all Braak-based ROIs were labeled T−, and any case in which regional positivity did not follow the hierarchical staging schema was labeled “discordant” and was excluded from tau PET and biological staging (n = 117 (3.5%); Figure 1B; Supplemental Tables 4-5). Individuals with hierarchical tau patterns were grouped into four tau PET stages: T− (tau negative across all Braak ROIs), T12+ (tau positive in only Braak I/II), T34+ (tau positive in Braak I-III or Braak I-IV), or T56+ (tau positive in Braak I-V or Braak I-VI). Among A+ individuals, tau PET stages were used to operationalize the 2024 Alzheimer’s disease biological stages^22^, as follows: Stage A: A+T−, Stage B: A+T12+, Stage C: A+T34+, and Stage D: A+T56+ (Figure 1B).

**Figure 1.**
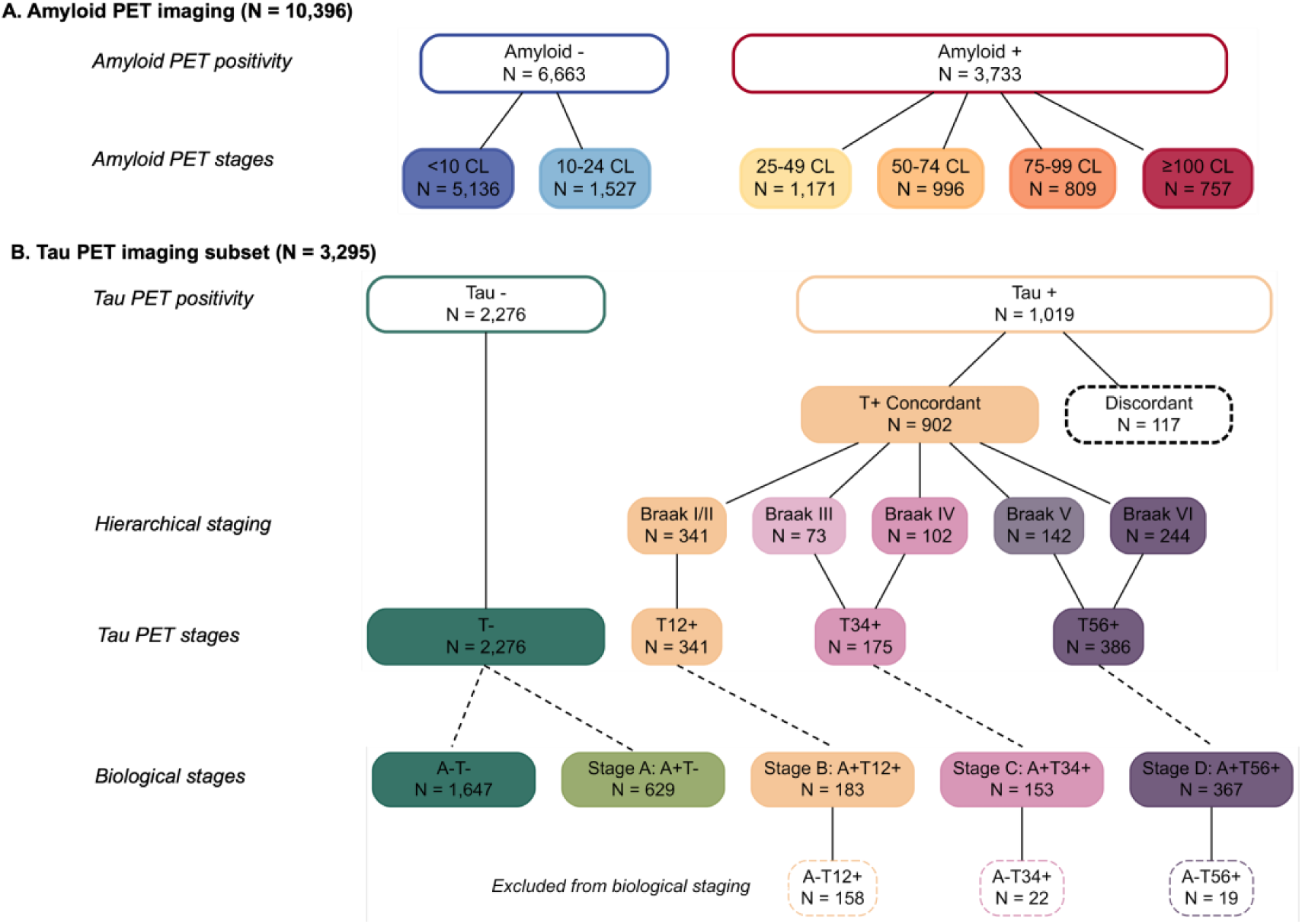
Amyloid and tau PET staging. Panel (**A**) shows the distribution of amyloid PET status, where positivity was determined as ≥25 CL, and amyloid PET staging, determined using six CL-based bins in the study sample (n=10,396). The breakdown of tau PET and biological staging for the subset of individuals from Panel A who also underwent tau PET imaging (n=3,295) is shown in panel (**B**). Tau positive individuals were assessed for concordance with Braak hierarchical staging. Those who followed the hierarchical staging were grouped into four hierarchical tau PET stages (e.g. T−, T12+, T34+, T56+). These tau PET stages were then used to operationalize the PET-based biological stages for AD (Jack et al. 2024), which require amyloid positivity for disease staging. Boxes with a dashed outline were excluded from staging schemas. Abbreviations: A+/−, Amyloid PET positivity; CL, centiloids; T+/−, Tau PET positivity

### Statistical Analyses

In parallel models, we examined the probability of dichotomous amyloid positivity (A+/−) and the cumulative probabilities of amyloid PET stages (6-level CL bins), tau PET stages, and Alzheimer’s disease biological stages according to age and clinical stage using logistic (binomial logit-link function) and ordinal logistic (cumulative logit-link function) regressions, respectively (VGAM and ordinal R packages). In the tau PET stage models, we examined the cumulative probabilities of tau stages according to CL in addition to age and clinical stage. For all logistic regression models, we covaried for cohort to account for differences in sampling and eligibility criteria. A forward selection method was used to test up to 2-way interactions between age (mean-centered) and clinical stage, and in the tau PET stage models up to 3-way interactions between age (mean-centered), CL (mean-centered), and clinical stage were examined. Interaction terms were retained if significant by the Wald statistical test (2-sided, *P*< .05). Estimated marginal probabilities and 95% confidence intervals (CIs) were used in figures. Odds ratios from logistic regression models are reported for each model term in the supplement. Statistical comparisons were reported at the mean age unless otherwise specified. Additionally, within each tau PET stage, differences in regional tau magnitude were examined across clinical stages using non-parametric Kruskal-Wallis tests, followed by post hoc Mann-Whitney U group-wise comparisons with Bonferroni correction. Finally, the alignment between Alzheimer’s disease biological stages and clinical stages was examined using the integrated approach described by Jack et al.^22^, where the expected disease progression was defined as biological Stages A and B in CU (“Clinical Stages 1-2”), Stage C in MCI (“Clinical Stage 3”), and Stage D in dementia (“Clinical Stages 4-6”).

## Results

### Participant characteristics

Among 10,396 participants with amyloid PET imaging, 5,948 (57%) were female with an average (SD) age of 71.9 (7.1) years (Table 1 and Supplementary Table 6).

### Amyloid PET stages across the clinical continuum

Overall, 36% (n = 3,733) of the amyloid PET sample were A+. As expected, the observed proportion of A+ increased with increasing clinical stage (Figure 2A; A+ CU, 28%; A+ MCI 50%; A+ Dementia 75%; Figure 2A). The median difference between the observed and predicted frequencies of A+ was 4.6% [IQR: 0.1% to 9.0%] across age bin comparisons indicating good model fit (Supplementary Tables 7-8). Predicted frequencies of A+ increased with clinical severity and age, though age had a weaker effect among those with dementia (clinical stage×age, *P*=.005; Figure 2B; Supplementary Tables 9-10). On average, the estimated frequency of A+ was twice as high in MCI compared to CU (CU: 25% (95% CI, 24%-27%); MCI: (51% (95% CI, 48%-54%)); Mean difference, 25% (95% CI, 22%-29%), *P*<.001) and about 1.5 times as likely in dementia compared to MCI (Dementia: 77% (95% CI, 75%-80%); Mean difference, 26% (95% CI, 23%-30%), *P*<.001).

**Figure 2.**
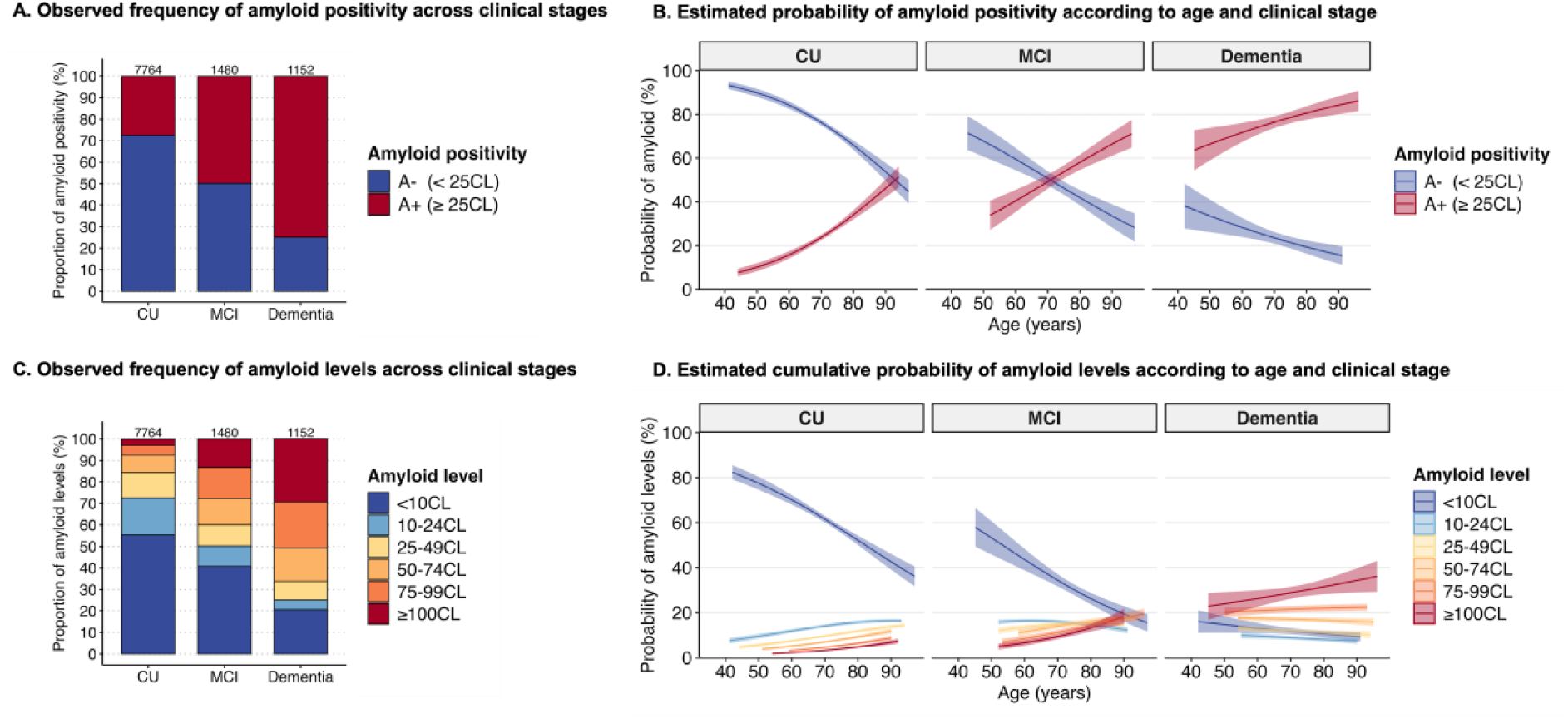
Frequency of amyloid PET severity across clinical stages. The observed frequency (**A**) and estimated probability (**B**) of amyloid status (e.g., amyloid positivity and negativity) is shown across clinical stages. In panel (**B**), the probability of amyloid status was modeled using age (*P*<.001), clinical stage (*P*<.001), cohort (*P*<.001), and age by clinical stage (*P*=.005) as predictors. The observed frequency (**C**) and estimated probability (**D**) of amyloid levels indicating amyloid severity is shown across clinical stages. In panel (**D**), the cumulative probability of amyloid levels was modeled using ordinal logistic regression with age (*P*<.001), clinical stage (*P*<.001), cohort (*P*<.001), and age by clinical stage (*P<*.001) as predictors. Shaded areas indicate the 95% confidence intervals for the probability estimates, which are shown for the observed age range for each biomarker group and clinical stage. Sample sizes of clinical stages are listed across the tops of the bars in panels (**A**) and (**C**). Abbreviations: *CL*, centiloid; *CU*, cognitively unimpaired; *MCI*, mild cognitive impairment.

CL-based amyloid staging showed higher amyloid levels were more common with increasing clinical severity; however, the full spectrum of amyloid severity was observed within each clinical stage (Figure 2C). Analyses estimating the cumulative probabilities of amyloid levels (Figure 2D) indicated a significant age×clinical stage interaction (*P* <.001), indicating that the age-related probability of elevated amyloid differed by clinical stage (Supplementary Tables 10-11). In CU, the frequency of amyloid levels of ≥10 CL increased with increasing age, while the frequency of amyloid levels <10 CL decreased with increasing age. On average, the lowest amyloid level (<10 CL) was most common in CU (52% (95% CI, 50%-53%)), followed by 10-24 CL (16% (95% CI, 15%-17%)) and 25-49 CL (12% (95% CI, 11%-13%)), and amyloid levels above 50 CL were less common, accounting for ∼ 20% of CU (50-74 CL (9% (95% CI, 9%-10%)); 75-99 CL (6% (95% CI, 6%-7%)); ≥100 CL (5% (95% CI, 4%-5%)); Figure 2D).

The age×MCI interaction was not significant (OR: 1.00; 95% CI: 0.99–1.01; *P* = .95), suggesting that the effect of age on amyloid levels is similar between CU and MCI. In MCI, the frequency of amyloid levels <25 CL (e.g. <10 CL and 10-24 CL) decreased with age, and estimates of amyloid levels ≥25 CL increased with age. In contrast, the age×dementia interaction was significant (OR: 0.97; 95% CI: 0.96–0.99; *P* = .001), indicating that in individuals with dementia, each additional year of age slightly reduces the odds of being in a higher amyloid level compared to CU. For example, at age 70, lower amyloid levels predominated in MCI, but by age 80, levels >10 CL were fairly evenly distributed. In dementia, age had a minimal effect, with the highest frequency observed at ≥100 CL (36% (95% CI, 33%-40%)) followed by 75-99 CL (23% (95% CI, 21%-24%)) and 50-74 CL (95% CI, 16% (15%-17%); Figure 2D).

### Tau PET stages across the amyloid and clinical continuums

Of the tau PET imaging subsample (n = 3,295; Supplemental Table 12), sixty-nine percent (n = 2,276) were T− across all Braak-based ROIs (Figure 1B). Among T+ individuals (n = 1,019), the majority (n = 902 (89%)) had tau PET spatial patterns concordant with the hierarchical patterns characterized by neuropathological studies and were included in the tau PET staging schema.^39^ Among those with hierarchical tau patterns, we examined the magnitude of regional tau burden within each tau PET stage across clinical stages (Supplemental Fig. 9). No differences in the magnitude or regional tau PET burden were observed across clinical diagnosis groups among T− or T12+ individuals. Among T34+ individuals, the magnitude of tau in Braak 3 and Braak 4 ROIs was significantly higher among MCI and dementia compared to CU. Among those with widespread tau T56+, a stepwise increase in the magnitude of tau in Braak 3,4, 5 and 6 ROIs was observed across clinical stages.

A total of 117 (11%) of T+ had quantitatively non-hierarchical “discordant” tau patterns and were therefore excluded from the tau PET staging schema and all subsequent analyses (Figure 1B). Visual review of the discordant tau cases revealed that 36% were visually tau negative (n = 42), while 22% showed visually hierarchical patterns concordant with Braak staging (n = 26; Supplemental Tables 4-5). The remaining 42% exhibited discordant tau positivity patterns which fell into two main categories: medial temporal lobe (MTL) sparing (i.e. sparing Braak 1-2; n = 34) and atypical uptake (sparing Braak 3 and/or Braak 4; n = 15)). Differences between quantitatively and visually assessed tau patterns within this group was attributed to a range of factors including off target binding (n = 25), threshold effects (n = 31), asymmetry (n = 24), atrophy (n = 4), and/or reference region uptake issues (n = 2). Discordant cases are detailed in Supplemental tables 4-5.

Observed frequencies of tau PET stages across amyloid levels and clinical stages is shown in Figure 3A. Of 2087 CU individuals, 339 (16%) were tau PET positive, of which most had tau isolated to the medial temporal lobe (T12+, n = 228 (67%)) and a third with tau spread outside the medial temporal lobe (T34+, n = 64 (19%); T56+, n = 47 (14%)). Notably, while T− (84%) was most common among CU, T12+ was observed across all amyloid levels and T34+ and T56+ were more common at amyloid levels >75 CLs (Figure 3A). Tau PET positivity was more frequent among individuals with MCI (273 out of 663 (41%)) and highest among those with dementia (290 out of 428 (68%)). Individuals with MCI had higher proportions of tau pathology at each amyloid level compared to CU individuals, such that at 50-74 CL, tau positivity (64% (11% T12+, 26% T34+, 27% T56+)) became more common than T− (36%). Individuals with dementia showed the most advanced tau stages, with a higher proportion of T56+ at all amyloid levels compared to CU and MCI. Overall, across clinical stages, tau severity was generally more common with higher CLs and worsening clinical stage; however, the full spectrum of tau severity was observed across all amyloid levels and clinical stages (Figure 3A).

**Figure 3.**
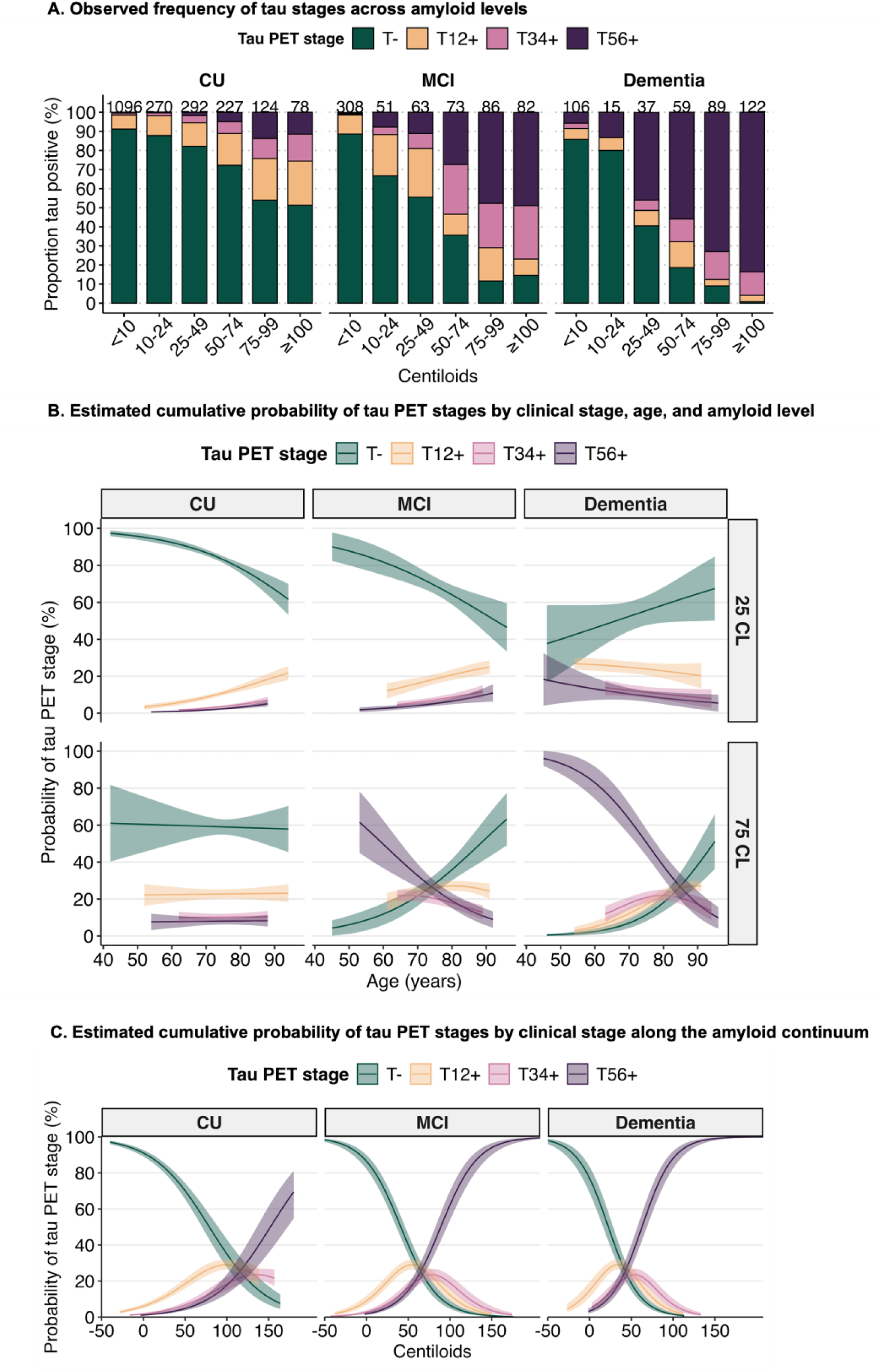
Frequency of tau PET severity across clinical stages. The observed frequency of hierarchical tau PET stages is shown across amyloid levels and by clinical stage in panel (**A**). Sample sizes for each subgroup are listed atop each bar. The cumulative probability of tau PET stages in shown in panels **(B**) and (**C**) and was modeled using ordinal logistic regression with centiloids (*P*<.001), age (*P*<.001), clinical stage (*P*<.001), cohort (*P*<.001), centiloids by clinical stage (*P*<.001), age by clinical stage (*P*<.001), centiloids by age (*P*<.001) and age by centiloids by clinical stage (*P*=.002) as predictors. Shaded areas indicating the 95% CIs for the probability estimates. Shaded areas indicating the 95% CIs for the probability estimates. Panel (**B**) details the age-related probability of various tau PET stages by age at tau PET scan across clinical stages (columns) at predicted low (25 CL) and high (75 CL) amyloid levels. In panel (**B**), probability curves reflect the observed age range for each tau PET stage and clinical stage. Panel (**C**) shows the amyloid-related probability of tau PET stages across clinical stages (columns) with probability curves shown for the observed centiloid range for each tau PET and clinical stage. Abbreviations: *CL*, centiloid; *CU*, cognitively unimpaired; *MCI*, mild cognitive impairment.

Ordinal logistic regression analyses estimating the cumulative probabilities of tau stages indicated a significant age×CL×clinical stage interaction, suggesting a nuanced pattern in how age and amyloid interact across clinical stages to influence tau severity (*P*<.001; Supplementary Tables 13-14). To probe this three-way interaction, we estimated age-related probabilities at two amyloid levels: low (25 CL) and high (75 CL; Figure 3B). At low amyloid levels, older age was associated with a higher likelihood of tau PET severity in CU (OR = 1.06, 95% CI (1.04, 1.08), *P*<.001) and in MCI (OR = 1.05, 95% CI (1.02, 1.08), *P*<.001), indicating 6% and 5% increase in odds of a higher tau PET stage per year of age in CU and MCI, respectively. By contrast, in dementia older age was not related to a higher likelihood of tau PET severity (OR = 0.97, 95% CI (0.94, 1.02), *P*=0.12). At higher amyloid levels, age effects shifted. Among CU individuals, there was no significant association between age and tau PET severity (OR = 1.00, 95% CI (0.97, 1.03), *P* = 0.85). However, among impaired individuals, older age was associated with lower odds of advanced tau PET stage in MCI (OR = 0.93, 95% CI (0.90, 0.96), *P* < .0001) and dementia (OR = 0.89, 95% CI (0.86, 0.92), *P* < .0001). For example, the likelihood of T56+ was highest in the presence of clinical impairment among those with younger ages and those with higher CLs (Figure 3B).

We next examined the average frequency of tau PET stages across the amyloid continuum at the average age at tau PET (∼72 years; Figure 3C). On average across clinical stages, the pattern of probability curves for each tau PET stage along the amyloid continuum were similar, such that the probability estimates of remaining T− decreased with increasing CLs, estimates for T12+ and T34+ peaked, and estimates of T56+ increased with increasing CLs (Figure 3C). While the peak amyloid-related probability for T12+ and T34+ was similar across all clinical stages (∼29% T12+, ∼24% T34+), the CL value at which the probability curves peaked for T12+ and T34+ differed considerably by clinical stage. On average, T12+ peaked earlier than T34+ and the peak CL value progressively decreased with increasing clinical severity (Peak T12+: CU 100 CL> MCI 54 CL > Dementia 34 CL; Peak T34+; CU 137 CL > MCI 78 CL > Dementia 54 CL; Supplementary Table 15).

### Alzheimer’s disease biological stages across the clinical continuum

PET-based Alzheimer’s disease biological stages included 1,332 A+ individuals with tau PET staging (Stage A: A+T−, n = 629; Stage B: A+T12+, n = 183; Stage C: A+T34+, n = 153; Stage D: A+T56+, n = 367) and 1,647 biomarker negative (A−T−) individuals were used as a reference group in subsequent analyses (Figure 1B). The observed frequencies of biological stages demonstrated increasing severity in biological stage with increasing severity in clinical stage; notably, however, each biological stage was observed across each clinical stage (Figure 4A).

**Figure 4.**
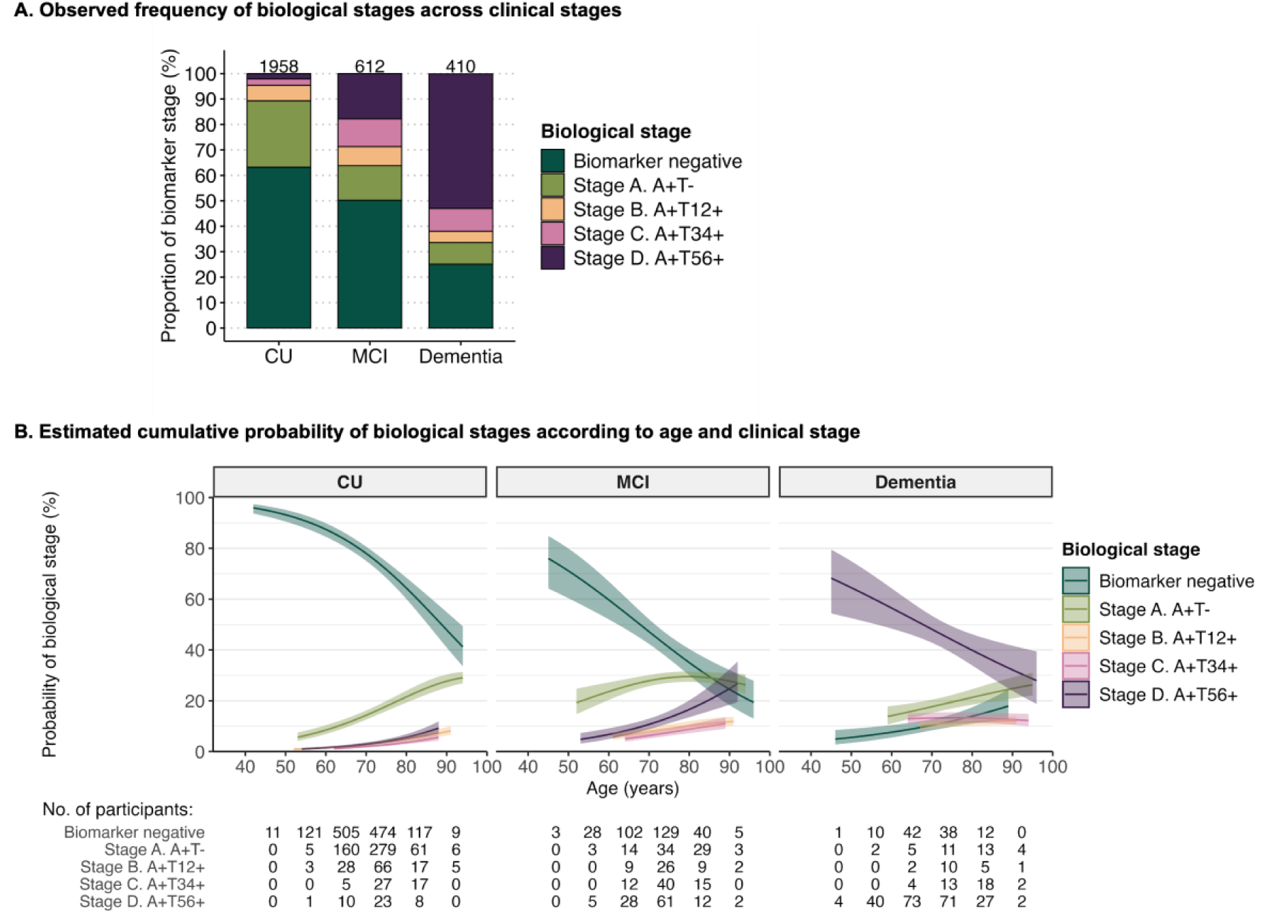
Frequency of Alzheimer’s disease biological stages across clinical stages. The observed frequency of PET-based Alzheimer’s disease biological stages is shown across clinical stages in panel (**A**). Sample sizes for each subgroup are listed on the top of each bar in (**A**). In panel (**B**) the probability of the biological stages is shown according to age and clinical stage. In panel (**B**) the estimated cumulative probability of biological stages was estimated using ordinal logistic regression with age (*P*<.001), clinical stage (*P*<.001), cohort (*P*<.001), and age by clinical stage (*P*<.001) as predictors. Shaded areas indicate the 95% confidence intervals for the probability estimates, which are shown for the observed age range for each biomarker group and clinical stage. Abbreviations: *CU*, cognitively unimpaired; *MCI*, mild cognitive impairment.

Ordinal logistic regression analyses indicated a significant age×clinical stage interaction (*P*<.001), suggesting that the age-related probability of biological stages differed by clinical stage (Figure 4B; Supplementary Tables 16-17). Among CU and MCI, the estimated frequencies of biological stages generally increased with older age compared to the biomarker negative reference group, indicating a higher likelihood of more severe biological stages with older age prior to dementia. By comparison, in dementia, the probability of Stage D decreased with age (age × dementia, OR = 0.90, 95% CI (0.88-0.93), *P*<.001), indicating that younger individuals were more likely to exhibit advanced tau pathology compared to older individuals. On average, tau positivity within biological stages increased with advancing clinical severity (CU<MCI<Dementia; Stage B, A+T12+ 3%<8%<11%; Stage C, A+T34+ 2%<7% <13%; Stage D, A+T56+ 3%<11%<47%) with the largest increase in probability observed in Stage D (Dementia vs MCI: Mean difference, 38% (95% CI, 32%-43%), *P*<.001; Dementia vs CU: Mean difference, 50% (95% CI, 45%-55%), *P*<.001).

Additionally, we implemented the integrated biological and clinical staging schema proposed in the recently revised criteria^22^ for diagnosis of Alzheimer’s disease in Figure 5. Generally, we observed agreement between biological and expected clinical stage, such that most individuals in biological stages A and B were CU, and those in stage D tended to have dementia. While biological and clinical stages broadly aligned (69%), notable discrepancies were observed. In Stages A-C, up to a third of individuals showed more advanced clinical symptoms than expected based on their biological stage, and in Stages C and D, ∼32% and 40% respectively, showed lower than expected clinical impairment based on their biological stage (Figure 5).

**Figure 5.**
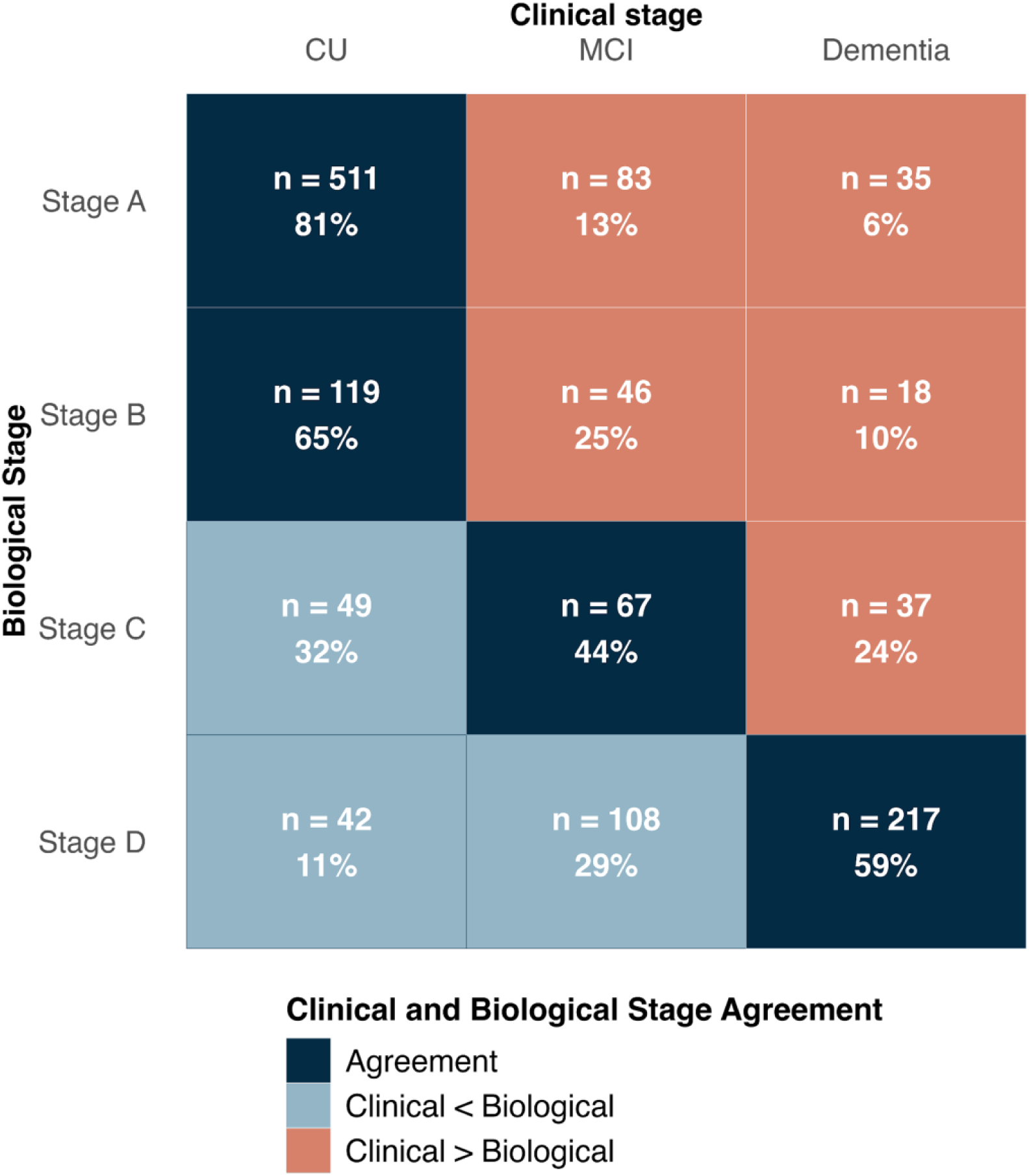
Integrated biological and clinical staging framework. Stacked tile plot showing the distribution of individuals by clinical stage (CU, MCI, Dementia; x-axis) and PET–based biological stage (Stages A–D; y-axis). Each cell is colored based on the alignment or disparity between clinical and biological staging using the integrated biological and clinical framework outlined by the Alzheimer’s Association (Dark blue = agreement; light blue = lower than expected clinical stage given biological stage; orange = higher than expected clinical stage given biological stage). Each cell shows the number of individuals (n) and the percentage of the total within each biological stage row. Abbreviations: *CU*, cognitively unimpaired; *MCI*, mild cognitive impairment.

## Discussion

In this study, we leveraged large-scale standardized PET data across multiple cohorts to stage amyloid and tau PET severity in individuals from Alzheimer’s disease research cohorts spanning the clinical continuum. Amyloid positivity and severity generally increased with clinical severity and age, though age-related effects were attenuated in dementia. Tau PET severity generally increased with amyloid and clinical severity, with widespread brain tau (T56+) most common in dementia among individuals with higher CLs and at younger ages. Notably, within each clinical stage the full continuum of amyloid and tau PET severity was observed, underscoring the presence of biological heterogeneity. Integrating amyloid and tau PET-based stages into Alzheimer’s disease biological stages^22^ revealed general agreement between biological and clinical stages, with approximately a third of individuals showing either worse or better than expected clinical function relative to their biological stage. Together, these findings emphasize the utility of PET-based staging in elucidating heterogeneity along the clinical disease continuum.

Our MRI-free PET processing approach allowed for the inclusion of PET data without a corresponding MRI and included individuals from established Alzheimer’s disease studies (WRAP, A4, ADNI) in conjunction with individuals from the NACC-ADRC program (SCAN-Mixed Protocol; SCAN-Prospective), which reflects Alzheimer’s disease and related disorders more broadly and includes a wider range of clinical presentations (Supplementary Table 6, Supplementary Methods). Although other work^26,28^ has leveraged pre-existing PET datasets to define disease stages based on amyloid and tau PET status, our current work is the first to standardize PET image processing and thresholding methods across cohorts before staging and characterizing PET severity. PET standardization included selecting a primary acquisition timing window for each ligand across cohorts and then subsequently using a standardized pipeline to extract SUVRs.^41^ As cohorts often differ in acquisition timing windows as well as image processing pipelines, both of which have been shown to influence the magnitude of SUVRs,^47,48^ ^49,50^ these initial PET standardization steps should provide more robust probability estimates of amyloid and tau PET-based stages reported herein.

Consistent with previous studies,^51,52^ we observed that, on average, the predicted probability of A+ increased with clinical severity (CU:28% < MCI:50% < Dementia:75%). Despite including individuals from the NACC ADRC program, who represent a broad spectrum of clinical presentations and non-Alzheimer’s disease syndromes^53^, our average estimates of amyloid positivity across clinical stages are nearly identical to previously published estimates.^51,52^ To evaluate the continuum of amyloid burden, we staged amyloid severity by dividing the CL scale, a common standardized measure of global brain amyloid, into six CL-based bins.^11^ We observed that in CU, the probability of amyloid ≥10CL increased with age, supporting the hypothesis that early or “sub-threshold” amyloid (e.g., 10-24 CL) may be a meaningful indicator of disease progression and highlighting the potential importance of “sub-threshold” amyloid for early intervention and primary prevention efforts.^54–56^ Amyloid ≥100 CL was rare in CU (∼5%), but nevertheless may represent an important subgroup for analyses geared towards mechanisms of resilience or imminent risk of progression. Compared to CU, individuals with MCI had more evenly distributed probabilities across amyloid levels, underscoring the importance of characterizing this group with amyloid PET to provide an indicator of whether underlying Alzheimer’s disease pathology may be driving impairment. Additionally, in MCI the cumulative probabilities of amyloid levels showed a marked shift with age, where after age ∼80 we observed a crossing of amyloid level probability curves, suggesting that individuals with MCI were more likely to transition from lower to higher amyloid levels with increasing age. By comparison, the average probability of the highest amyloid level was predominant in dementia (e.g., ≥100 CL, 36%, 95% CI (33%-40%)), suggesting that most individuals with dementia that enroll into the included cohorts tend to have the highest CLs, regardless of age. Finally, the continuum of amyloid severity observed within and across each clinical stage underscores the heterogeneity in amyloid burden across the clinical continuum.

Another goal of the current work was to characterize the frequency of tau PET severity as a function of amyloid, age, and clinical stage. Although there is no consensus on the best method for quantifying tau PET severity, approaches that incorporate tau spatial distribution have consistently shown an increasing risk of clinical impairment with greater tau spread.^19,20,57,58^ Correspondingly, we used a Braak-based hierarchical staging approach to assess tau PET severity based on spatial extent of tau positivity. Consistent with previous studies^18–21^, we found that 97% of our tau PET sample were either tau negative or exhibited hierarchical spatial patterns aligning with Braak staging. Notably, within later hierarchical tau PET stages (T34+ and T56+), we observed differences in the magnitude of regional tau burden across clinical stages (Supplemental Fig. 9). Among individuals with severe tau (T56+), we observed a stepwise increase in tau burden with advancing clinical stage in tau PET ROIs corresponding to Braak 3, 4, 5 and 6—highlighting additional variability in tau PET magnitude, even among those with widespread tau uptake. Integrating both the spatial extent and magnitude of tau PET signal may enhance frameworks for staging tau pathology and tracking disease progression. However, it is also possible that these two dimensions provide distinct information and should be evaluated separately to assess disease progression or therapeutic response. As tau PET imaging becomes more widely adopted, it will be important to clarify how each metric—independently or in combination—can inform clinical trial design and guide treatment decisions.

While implementing a hierarchical tau staging schema allowed us to quantitatively assess both the severity and spread of tau, it also excluded 3% of cases with non-hierarchical tau patterns. Despite quantitatively discordant patterns of tau positivity, visual inspection of these images revealed that most were either tau negative or had visually concordant tau patterns, where discordance was due to threshold effects, asymmetry, off target binding or reference region issues. Visual inspection also revealed two biologically plausible non-hierarchical patterns of tau positivity: MTL sparing (n = 34) and atypical (sparing Braak 3 and/or Braak 4; n = 15) patterns. Although discordant cases were excluded to preserve the quantitative approach and fidelity of the spatial extent-based staging framework, the MTL sparing and atypical patterns have been extensively studied^59–61^ and may represent clinically and/or biologically meaningful subtypes. Future staging efforts aimed at capturing the full spectrum of tau PET patterns—including those that fall outside of canonical Braak staging— will be critical for understanding atypical Alzheimer’s disease phenotypes, improving classification accuracy, and ensuring inclusivity in biomarker-based models of disease progression.

Overall, we observed complex relationships between the frequency of tau PET stages and age, amyloid, and clinical stage. In analyses estimating the cumulative probabilities of tau PET stages, we observed that on average, within each clinical stage, the CL value at which T34+ probability peaked was significantly higher than for T12+ (Supplemental Table 15), suggesting that more advanced tau stages and wider spatial extent generally requires more amyloid accumulation—a finding consistently observed across tau PET studies.^13,19–21^ While the spatial extent and severity of tau generally increased with higher amyloid across clinical stages, within a given amyloid level (e.g., 50-74 CL) all tau stages were observed. Further, the probabilities of T12+ and T34+ peaked at progressively lower CLs as clinical severity increased (e.g., CU > MCI > Dementia; Figure 3B, Supplementary Table 15), suggesting that factors beyond amyloid, like co-morbidities or co-pathologies, may be contributing to impairment among those with MCI and dementia which may reduce the amount of amyloid needed for tau propagation.^62^ Jointly, these findings align with previous studies^21,25,28,31,61,63,64^ and highlight that heterogeneity in tau PET within and across clinical stages remains after accounting for overall amyloid burden.

We also found that age moderated the effects of amyloid and clinical stage on the cumulative probability of tau severity, whereby late-stage tau (T56+) was most prominent in dementia at younger ages and higher amyloid levels. Similar age-related patterns of tau severity across clinical stages emerged in the biological stage frequency analyses, where older CU and MCI A+ participants were more likely to progress to advanced biological stages, while younger individuals with dementia had a higher likelihood of severe tau (e.g., Stage D A+T56+). The decreasing trend of severe tau (both T56+ and A+T56+) with age is consistent with recent PET and neuropathological studies^30,65^ and suggests that those with higher tau levels may develop dementia earlier. Although mechanisms driving an increased risk of widespread brain tau among A+ individuals remain unclear, younger-onset Alzheimer’s disease cases^66^ are known to exhibit higher levels of cortical tau, and multiple studies have demonstrated an inverse association between tau burden and age in Alzheimer’s disease cohorts.^30,31,65,67^ Additionally, greater synaptic density and/or neuronal connectivity in younger individuals may facilitate tau propagation.^68^ Older individuals with dementia and advanced tau pathology may be underrepresented in research cohorts; thus, longitudinal studies across the aging continuum are needed to better understand relationships between age and tau PET severity.^69^

Among CU individuals, we observed that 16% had elevated tau, of which most had tau confined to the medial temporal lobe (T12+ (67%)) and a small but significant proportion had tau spread outside the medial temporal lobe (T34+ (19%) and T56+ (13%)). Consistent with previous studies^65,70^, tau positivity occurred across the full range of amyloid levels, indicating that preclinical tau can emerge even in the absence of amyloid. By contrast, the observed frequency of advanced tau stages (e.g., T34+ and T56+) in CU was lower, but more common among those with higher amyloid levels (>75 CL), suggesting a threshold above which amyloid burden is associated with greater tau spread in the preclinical stage. These findings highlight the heterogeneity of tau pathology in CU individuals and reinforce the idea that tau PET can detect preclinical disease progression, especially in the context of elevated amyloid burden. The presence of T12+ across all amyloid levels suggests that early tau accumulation in the medial temporal lobe may be part of aging-related changes, not necessarily Alzheimer’s disease-specific, whereas tau spread outside the medial temporal lobe (e.g., T34+ and T56+) appears to be more tightly linked to amyloid accumulation and may reflect a transition toward symptomatic Alzheimer’s disease. Correspondingly, these T34+ and T56+ CU individuals with elevated amyloid may represent a critical window for early intervention, particularly for anti-tau or combination therapies. These findings also underscore the need for longitudinal follow-up to determine which patterns of tau positivity predict clinical progression and which remain stable over time, refining risk prediction models and informing therapeutic timing.

Given the growing integration of PET imaging into the field of Alzheimer’s disease research, amyloid and tau PET have become central components of the biological staging criteria for AD.^22^ In the current study, we applied these PET-based stages across multiple cohorts and tracers and found that biological stage severity generally increased with clinical stage severity, aligning with established models of Alzheimer’s disease progression. Our operationalization of biological stages C and D were based on spatial extent of tau; nevertheless, our results largely align with a recent study^71^ that used tau magnitude to distinguish these stages. Across both approaches, there was strong concordance between biological and clinical stages, such that most individuals in biological stages A and B were CU, and those in stages C and D tended to have MCI or dementia. However, approximately one-third of participants showed discordance between biological and clinical stage: 17% of individuals exhibited more advanced clinical symptoms than expected and 15% showed less severe clinical impairment relative to their biological stage. By design, discordance within this integrated biological and clinical staging framework highlights the individual variability in the relationship between pathology and clinical symptoms and captures both typical and atypical patterns of disease manifestation.^22^ Continued investigation of these biological and clinical stage mismatch cases may offer important clues for the evolving precision medicine approach to Alzheimer’s disease and related disorders. Individual variability in biological and/or clinical progression may stem from differences in co-pathologies^72^, co-morbidities, cognitive resilience, or cohort sampling; thus, future work is needed to understand the clinical relevance of PET-based Alzheimer’s disease biological stages, especially where biological and clinical stages diverge.

### Strengths and Limitations

The strengths of this study include standardizing PET imaging across multiple cohorts and PET tracers to implement and characterize amyloid and tau PET-based staging frameworks in individuals spanning the clinical continuum. While this is one of the first attempts to operationalize PET-based biological staging criteria using standardized imaging measures and thresholding methods, individuals in this study were enrolled in Alzheimer’s disease research cohorts that were not randomly sampled from the community; therefore, the probability estimates reported herein do not reflect the broader U.S. population. Further, across all models of amyloid and tau PET severity, we observed non-parallel effects across cohorts highlighting systematic differences in PET severity across cohorts and potential variability due to sampling or other unmeasured confounders. Additional work is needed to characterize the impact of cohort sampling on the frequency of PET severity in Alzheimer’s disease. Also, much of the sample was tau negative and the sample sizes for tau positive PET and biological stages, and those aged younger than 50 and older than 90 years were relatively small and should be interpreted with caution. Finally, given the cross-sectional nature of the current study, longitudinal follow-up will be necessary to fully characterize the associations of amyloid and tau PET-based stages with clinical progression over time.

## Conclusions

By leveraging large-scale standardized PET data from multiple cohorts, we provide robust probability estimates of amyloid and tau PET severity along the clinical continuum. Together, our findings highlight heterogeneity in disease progression and the potential for amyloid and tau PET-based staging to identify windows for therapeutic intervention and enhance understanding of disease progression.

## Data availability

Data used in this manuscript are available by request at the ADNI data repository at the Laboratory of Neuroimaging (http://adni.loni.usc.edu.), the National Alzheimer’s Coordinating Center (https://naccdata.org/), and through the National Institute on Aging Genetics of Alzheimer’s Disease Data Storage Site (https://dss.niagads.org/datasets/ng00067/).

## Acknowledgements

The Alzheimer’s Disease Sequencing Project Phenotype Harmonization Consortium (ADSP-PHC) is funded by NIA (U24 AG074855, U01 AG068057 and R01 AG059716). Biological samples and associated phenotypic data used in primary data analyses were stored at Study Investigators institutions, and at the National Centralized Repository for Alzheimer’s Disease and Related Dementias (NCRAD, U24AG021886) at Indiana University funded by NIA. Associated Phenotypic Data used in primary and secondary data analyses were provided by Study Investigators, the NIA funded Alzheimer’s Disease Centers (ADCs), and the National Alzheimer’s Coordinating Center (NACC, U24AG072122) and the National Institute on Aging Genetics of Alzheimer’s Disease Data Storage Site (NIAGADS, U24AG041689) at the University of Pennsylvania, funded by NIA. Harmonized phenotypes were provided by the ADSP Phenotype Harmonization Consortium (ADSP-PHC), funded by NIA (U24 AG074855, U01 AG068057 and R01 AG059716) and Ultrascale Machine Learning to Empower Discovery in Alzheimer’s Disease Biobanks (AI4AD, U01 AG068057). This research was supported in part by the Intramural Research Program of the National Institutes of health, National Library of Medicine. Contributors to the Genetic Analysis Data included Study Investigators on projects that were individually funded by NIA, and other NIH institutes, and by private U.S. organizations, or foreign governmental or nongovernmental organizations. Data used in preparation of the present article were obtained from the A4 database (https://www.a4studydata.org/), the ADNI database (adni.loni.usc.edu), the NACC database (https://naccdata.org/requesting-data/nacc-data), and from the University of Wisconsin (https://wrap.wisc.edu/). Our deepest gratitude to all research participants for participation.

The Anti-Amyloid Treatment in Asymptomatic Alzheimer’s study (A4 Study) is a secondary prevention trial in preclinical Alzheimer’s disease, aiming to slow cognitive decline associated with brain amyloid accumulation in clinically normal older individuals. The A4 Study is funded by a public-private-philanthropic partnership, including funding from the National Institutes of Health-National Institute on Aging, Eli Lilly and Company, Alzheimer’s Association, Accelerating Medicines Partnership, GHR Foundation, an anonymous foundation and additional private donors, with in-kind support from Avid and Cogstate. The companion observational Longitudinal Evaluation of Amyloid Risk and Neurodegeneration (LEARN) Study is funded by the Alzheimer’s Association and GHR Foundation. The A4 and LEARN Studies are led by Dr. Reisa Sperling at Brigham and Women’s Hospital, Harvard Medical School and Dr. Paul Aisen at the Alzheimer’s Therapeutic Research Institute (ATRI), University of Southern California. The A4 and LEARN Studies are coordinated by ATRI at the University of Southern California, and the data are made available through the Laboratory for Neuro Imaging at the University of Southern California. The participants screening for the A4 Study provided permission to share their de-identified data in order to advance the quest to find a successful treatment for Alzheimer’s disease. We would like to acknowledge the dedication of all the participants, the site personnel, and all of the partnership team members who continue to make the A4 and LEARN Studies possible. The complete A4 Study Team list is available on: a4study.org/a4-study-team.

Data collection and sharing for this project was also funded by the Alzheimer’s Disease Neuroimaging Initiative (ADNI) (National Institutes of Health Grant U01 AG024904) and DOD ADNI (Department of Defense award number W81XWH-12-2-0012). ADNI is funded by the National Institute on Aging, the National Institute of Biomedical Imaging and Bioengineering, and through generous contributions from the following: AbbVie, Alzheimer’s Association; Alzheimer’s Drug Discovery Foundation; Araclon Biotech; BioClinica, Inc.; Biogen; Bristol-Myers Squibb Company; CereSpir, Inc.; Cogstate; Eisai Inc.; Elan Pharmaceuticals, Inc.; Eli Lilly and Company; EuroImmun; F. Hoffmann-La Roche Ltd and its affiliated company Genentech, Inc.; Fujirebio; GE Healthcare; IXICO Ltd.;Janssen Alzheimer Immunotherapy Research & Development, LLC.; Johnson & Johnson Pharmaceutical Research & Development LLC.; Lumosity; Lundbeck; Merck & Co., Inc.;Meso Scale Diagnostics, LLC.; NeuroRx Research; Neurotrack Technologies; Novartis Pharmaceuticals Corporation; Pfizer Inc.; Piramal Imaging; Servier; Takeda Pharmaceutical Company; and Transition Therapeutics. The Canadian Institutes of Health Research is providing funds to support ADNI clinical sites in Canada. Private sector contributions are facilitated by the Foundation for the National Institutes of Health (www.fnih.org). The grantee organization is the Northern California Institute for Research and Education, and the study is coordinated by the Alzheimer’s Therapeutic Research Institute at the University of Southern California. ADNI data are disseminated by the Laboratory for Neuro Imaging at the University of Southern California. A complete listing of ADNI investigators can be found at: http://adni.loni.usc.edu/wp-content/uploads/how_to_apply/ADNI_Acknowledgement_List.pdf.

We also acknowledge all the National Alzheimer’s Coordinating Center (NACC) ADRC research participants and the ADRC sites that provided PET data for this effort. The National Alzheimer’s Coordinating Center (NACC) database is funded by NIA/NIH Grant U24 AG072122. SCAN is a multi-institutional project that was funded as a U24 grant (AG067418) by the National Institute on Aging in May 2020. Data collected by SCAN and shared by NACC are contributed by the NIA-funded ADRCs as follows: Arizona Alzheimer’s Center - P30 AG072980 (PI: Eric Reiman, MD); R01 AG069453 (PI: Eric Reiman (contact), MD); P30 AG019610 (PI: Eric Reiman, MD); and the State of Arizona which provided additional funding supporting our center; Boston University - P30 AG013846 (PI Neil Kowall MD); Cleveland ADRC - P30 AG062428 (James Leverenz, MD); Cleveland Clinic, Las Vegas – P20AG068053; Columbia - P50 AG008702 (PI Scott Small MD); Duke/UNC ADRC – P30 AG072958; Emory University - P30AG066511 (PI Levey Allan, MD, PhD); Indiana University - R01 AG19771 (PI Andrew Saykin, PsyD); P30 AG10133 (PI Andrew Saykin, PsyD); P30 AG072976 (PI Andrew Saykin, PsyD); R01 AG061788 (PI Shannon Risacher, PhD); R01 AG053993 (PI Yu-Chien Wu, MD, PhD); U01 AG057195 (PI Liana Apostolova, MD); U19 AG063911 (PI Bradley Boeve, MD); and the Indiana University Department of Radiology and Imaging Sciences; Johns Hopkins - P30 AG066507 (PI Marilyn Albert, Phd.); Mayo Clinic - P50 AG016574 (PI Ronald Petersen MD PhD); Mount Sinai - P30 AG066514 (PI Mary Sano, PhD); R01 AG054110 (PI Trey Hedden, PhD); R01 AG053509 (PI Trey Hedden, PhD); New York University - P30AG066512-01S2 (PI Thomas Wisniewski, MD); R01AG056031 (PI Ricardo Osorio, MD); R01AG056531 (PIs Ricardo Osorio, MD; Girardin Jean-Louis, PhD); Northwestern University - P30 AG013854 (PI Robert Vassar PhD); R01 AG045571 (PI Emily Rogalski, PhD); R56 AG045571, (PI Emily Rogalski, PhD); R01 AG067781, (PI Emily Rogalski, PhD); U19 AG073153, (PI Emily Rogalski, PhD); R01 DC008552, (M.-Marsel Mesulam, MD); R01 AG077444, (PIs M.-Marsel Mesulam, MD, Emily Rogalski, PhD); R01 NS075075 (PI Emily Rogalski, PhD); R01 AG056258 (PI Emily Rogalski, PhD); Oregon Health and Science University - P30 AG008017 (PI Jeffrey Kaye MD); R56 AG074321 (PI Jeffrey Kaye, MD); Rush University - P30 AG010161 (PI David Bennett MD); Stanford – P30AG066515; P50 AG047366 (PI Victor Henderson MD MS); University of Alabama, Birmingham – P20; University of California, Davis - P30 AG10129 (PI Charles DeCarli, MD); P30 AG072972 (PI Charles DeCarli, MD); University of California, Irvine - P50 AG016573 (PI Frank LaFerla PhD); University of California, San Diego - P30AG062429 (PI James Brewer, MD, PhD); University of California, San Francisco - P30 AG062422 (Rabinovici, Gil D., MD); University of Kansas - P30 AG035982 (Russell Swerdlow, MD); University of Kentucky - P30 AG028283-15S1 (PIs Linda Van Eldik, PhD and Brian Gold, PhD); University of Michigan ADRC - P30AG053760 (PI Henry Paulson, MD, PhD) P30AG072931 (PI Henry Paulson, MD, PhD) Cure Alzheimer’s Fund 200775 - (PI Henry Paulson, MD, PhD) U19 NS120384 (PI Charles DeCarli, MD, University of Michigan Site PI Henry Paulson, MD, PhD) R01 AG068338 (MPI Bruno Giordani, PhD, Carol Persad, PhD, Yi Murphey, PhD) S10OD026738-01 (PI Douglas Noll, PhD) R01 AG058724 (PI Benjamin Hampstead, PhD) R35 AG072262 (PI Benjamin Hampstead, PhD) W81XWH2110743 (PI Benjamin Hampstead, PhD) R01 AG073235 (PI Nancy Chiaravalloti, University of Michigan Site PI Benjamin Hampstead, PhD) 1I01RX001534 (PI Benjamin Hampstead, PhD) IRX001381 (PI Benjamin Hampstead, PhD); University of New Mexico - P20 AG068077 (Gary Rosenberg, MD); University of Pennsylvania - State of PA project 2019NF4100087335 (PI David Wolk, MD); Rooney Family Research Fund (PI David Wolk, MD); R01 AG055005 (PI David Wolk, MD); University of Pittsburgh - P50 AG005133 (PI Oscar Lopez MD); University of Southern California - P50 AG005142 (PI Helena Chui MD); University of Washington - P50 AG005136 (PI Thomas Grabowski MD); University of Wisconsin - P50 AG033514 (PI Sanjay Asthana MD FRCP); Vanderbilt University – P20 AG068082; Wake Forest - P30AG072947 (PI Suzanne Craft, PhD); Washington University, St. Louis - P01 AG03991 (PI John Morris MD); P01 AG026276 (PI John Morris MD); P20 MH071616 (PI Dan Marcus); P30 AG066444 (PI John Morris MD); P30 NS098577 (PI Dan Marcus); R01 AG021910 (PI Randy Buckner); R01 AG043434 (PI Catherine Roe); R01 EB009352 (PI Dan Marcus); UL1 TR000448 (PI Brad Evanoff); U24 RR021382 (PI Bruce Rosen); Avid Radiopharmaceuticals / Eli Lilly; Yale - P50 AG047270 (PI Stephen Strittmatter MD PhD); R01AG052560 (MPI: Christopher van Dyck, MD; Richard Carson, PhD); R01AG062276 (PI: Christopher van Dyck, MD); 1Florida - P30AG066506-03 (PI Glenn Smith, PhD); P50 AG047266 (PI Todd Golde MD PhD).

Data used in preparation of this article were also obtained from the Wisconsin Registry for Alzheimer’s Prevention (WRAP: R01 AG021155). The WRAP is led by principal investigator Sterling C. Johnson, PhD at the University of Wisconsin Madison. We thank all the WRAP research participants that provided PET data for this effort.

## Funding

This work was supported by the NIH (U24 AG067418; U24 AG074855; U01AG082350; K01AG078443; K76-AG088554; K99AG071837) and the Alzheimer’s Association (AARFD-21-849349; AACSF-24-1307411). Additionally, this work was supported by funding from the Webb Family Foundation. The content is solely the responsibility of the authors and does not necessarily represent the official views of the National Institutes of Health.

## Competing interests

Dr. Mormino is a consultant to Neurotrack, Eli Lilly, and Roche and receives funding from NIH, Simons Foundation, Alzheimer’s Association, and Webb Family Foundation. All other coauthors report no disclosures.

## Supplementary material

Supplementary material is available at *Brain* online

