## Supplementary Methods for "Characterizing amyloid and tau PET-based stages across the clinical continuum"

**Supplementary Online Content**

<u>**Table of contents**</u>


***Supplementary Methods***

***Supplementary Figures***
**Figure 1.** Inclusion and exclusion criteria for amyloid and tau PET samples
**Figure 2.** Tau PET templates
**Figure 3.** Tau PET regions of interest
**Figure 4.** GMM BIC comparison: FTP
**Figure 5**. GMM BIC comparison: MK
**Figure 6.** Reclassification of GMM tau negative cases
**Figure 7.** GMM reclassified distributions: FTP scans
**Figure 8.** GMM reclassified distributions: MK scans
**Figure 9**. Regional tau PET SUVR Z-scores across clinical stages by hierarchical tau PET stage

***Supplementary Tables***
**Table 1**. PET timing windows and radiotracers
**Table 2**. Centiloid equations for MRI-Free amyloid PET scans
**Table 3.** Tracer-specific GMM: Summary of optimal models
**Table 4**. Summary of discordant tau PET positivity patterns
**Table 5.** Characteristics of participants with discordant tau patterns
**Table 6.** Amyloid PET imaging sample participant characteristics by cohort
**Table 7.** Observed probabilities of amyloid positivity
**Table 8.** Predicted probabilities of amyloid positivity
**Table 9.** Odds ratios from ordinal logistic regression predicting amyloid positivity
**Table 10.** Logistic model comparisons for amyloid outcomes
**Table 11.** Odds ratios from ordinal logistic regression predicting amyloid levels
**Table 12**. Tau PET imaging sample participant characteristics by cohort
**Table 13.** Logistic model comparisons for tau stage outcomes
**Table 14.** Odds ratios from ordinal logistic regression predicting tau stages
**Table 15.** Estimated centiloids at which the T12 and T34 probability curves peak across clinical stages
**Table 16.** Logistic model comparisons for biological stage outcomes
**Table 17.** Odds ratios from ordinal logistic regression predicting biological stages

***References***

***Supplemental Methods***

### Cohort descriptions

*ADNI*

The Alzheimer's Disease Neuroimaging Initiative (ADNI) is a multi-site longitudinal study designed to validate biomarkers for use in Alzheimer's disease clinical trials.[1] Information on ADNI enrollment, study design, and cohort composition has been described previously.[2-6] All data generated by the ADNI study are available via the Laboratory of Neuroimaging (LONI), an online data repository. Clinical stage was established from ADNI MERGE (dxsum.rdata) using the "DIAGNOSIS" variable. Participants in ADNI with an amyloid positron emission tomography (PET) scan and a diagnosis of cognitively unimpaired (CU), mild cognitive impairment (MCI), or dementia were included in the current study. Criteria for study enrollment of MCI individuals into ADNI has varied slightly between phases of the study and generally follows criteria for a single-domain amnestic presentation.[7] Dementia participants enrolled in ADNI have clinical presentations consistent with AD dementia with other neurodegenerative diseases excluded at baseline enrollment.

Within ADNI, 18F-florbetapir (FBP), 18F-florbetaben (FBB), and 11C-Pittsburgh Compound B (PIB) were available for amyloid PET. 18F-Flortaucipir (FTP) was used for tau PET. Summed PET images with a series description of "* Coreg, Avg, Std Img and Vox Siz, Uniform *Res*" were downloaded from LONI on 05/01/2024 and processed.

*A4*

The Anti-Amyloid Treatment in Asymptomatic Alzheimer disease (A4) Study is a prevention trial in CU older individuals aged 65–85 with evidence of elevated brain amyloid and took place across 67 sites in the US, Canada, Australia, and Japan (https://clinicaltrials.gov/ct2/show/ NCT02008357). Detailed inclusion and exclusion criteria for the A4 study has been described previously.[8] A4 amyloid scans and study data from the full screening dataset were downloaded from LONI on 01/18/2023. To receive an amyloid PET scan, individuals were determined to be clinically normal with the following criteria: 0 on the global Clinical Dementia Rating scale, 25–30 on the Mini-Mental State Examination, and 6–18 on the Logical Memory II test. All CU participants who were screened with amyloid PET for inclusion in the A4 study were included in our current analyses.

Within A4, FBP was used for amyloid PET and FTP for tau PET. Frame data for both amyloid and tau PET data were downloaded from LONI on 01/18/2023 and processed. FBP contained 4 five-minute frames corresponding to 50-70 minutes post-injection. All four frames were realigned and summed. FTP PET data contained 6 five-minute frames corresponding to 80-110 minutes post-injection. The first 4 frames corresponding to 80-100 minutes were selected, realigned, and summed.

*ADRC-NACC*

The National Alzheimer's Coordinating Center (NACC) is a data repository for the National Institute of Aging's Alzheimer's Disease Research Center (ADRC) program. Standardized data across ADRCs enrolled in NACC is collected via the Uniform Data Set (UDS) and submitted to NACC for storage and dissemination. NACC ADRC participants are currently drawn from 33 ADRCs and 4 exploratory centers across the United States (https://naccdata.org/). Although inclusion and exclusion criteria across the ADRC program is site specific and contains the full spectrum of Alzheimer's disease and related disorders (ADRD), the majority of ADRC enrollees with clinical impairment are suspected of having AD (https://naccdata.org/requesting-data/data-summary/uds).

Prior to 2021, NACC ADRC imaging was conducted through investigator led project and followed site-specific "mixed protocols." In January 2021, the SCAN initiative was implemented to standardize imaging acquisition protocols and analysis pipelines for ADRC PET and MRI and allow for harmonization of imaging data with other studies, including ADNI. Thus, the ADRC program currently encompasses two phases of PET imaging: Mixed Protocol, consisting of PET scans acquired using site-specific mixed protocols prior to the launch of SCAN, and SCAN, consisting of imaging data collected after January 2021 using standardized workflows. It is noteworthy that these older Mixed Protocol PET images did in fact follow acquisition protocols that for the most part are very similar to the SCAN protocol but were collected before centralized scanner qualification and thus followed site-specific reconstruction parameters and were not processed by the SCAN PET Core.

Within the Mixed Protocol sub-cohort, FBP, FBB, and PIB were available for amyloid PET. MK6240 and FTP were available for tau PET. These data were downloaded from either NACC or LONI between 2021 and 2025. Since this dataset did not involve standardized protocols, input data varied across individual NACC sites. 775 of 929 amyloid scans contained dynamic frame data. For this subset, the frames corresponding to each ligands recommended window were selected, realigned, and summed. The remaining 154 scans were received as summed images, which were used as inputs for processing. Within the SCAN sub-cohort, FBP, FBB, PIB, and NAV were available for amyloid PET. FTP and MK6240 were available for tau PET. Summed PET images with a series description of "* Coreg, Avg, Std Img and Vox Siz, Uniform *Res*" on LONI that underwent pre-processing by the SCAN PET Core were downloaded from LONI on 03/31/2025 and included in the current analysis.[9]

Study data for all NACC participants (including both Mixed Protocol and SCAN) were downloaded from NACC on 03/31/2025 (investigator_nacc69.csv). The breakdown of NACC participant data across Mixed Protocol and SCAN PET sub-cohorts in the main manuscript analyses is detailed in Tables 4-5). It is noteworthy that 246 amyloid scans from NACC were either not included in investigator_nacc69.csv or were missing age or clinical data (Figure 1A). This is likely due to different cadences of data release for clinical and PET data. Clinical stage for all NACC ADRC participants was determined using the "NACCUDSD" variable from the investigator_nacc69.csv and included normal cognition, Impaired-Not MCI, MCI, or Dementia. For the current study, those who were diagnosed as Impaired-Other (n=61) were combined with MCI.

*WRAP*

The Wisconsin Registry for Alzheimer's Prevention (WRAP) cohort is a risk-enriched longitudinal study designed to identify mid-life factors associated with the progression of Alzheimer's disease.[10] WRAP enrollment and assessment began in 2001, enrolling cognitively unimpaired individuals aged 40–65. WRAP participants undergo biennial cognitive, health, and optional biomarker assessments. Imaging and meta-data including a consensus diagnosis were downloaded from LONI on 10/22/2024. WRAP participants with available amyloid PET imaging and a diagnosis of CU-declining, CU-stable, Impaired-Other, MCI, or dementia were included in the current study. CU-declining and CU-stable were combined into the CU group, and Impaired-Other (n=8) was combined with MCI into the MCI group.

Within WRAP, PIB and NAV were used for amyloid PET and MK6240 for tau PET. Frame data for both amyloid and tau PET data were downloaded from LONI on 10/22/24 and processed. PIB data contained 17 frames corresponding to 0-70 minutes post-injection. The last four five-minute PIB frames corresponding to 50-70 minutes post-injection were realigned and summed. NAV data contained 4 frames corresponding to 50-70 min post-injection. All frames were realigned and summed. MK6240 PET data contained 8 frames corresponding to 70-110 minutes post-injection. The last 4 frames corresponding to 90-110 minutes were selected, realigned, and summed.

**Image Processing: Quality control**

*Pre-processing PET Quality Control (QC): QC Timing*

Across all cohorts (A4, ADNI, WRAP, SCAN and Mixed Protocol), acquisition timing information was used to only include data within 10-minutes before or after the designated recommended acquisition start time for each (Table 1). The one exception was PIB, in which the majority of scans were collected 50-70 minutes post-injection (n=645 SCAN, n=920 Mixed Protocol, n=1197 WRAP, and n=228 ADNI). However, a subset of n=433 PIB scans from SCAN was collected between 40-60 minutes post-injection and was adjusted to a 50-70 minute scale using a linear regression (Table 1). Limiting data to a single acquisition timing window per tracer resulted in the exclusion of 114 amyloid scans and 6 tau scans (see QC timing fails, Figure 1AB).

*Post-processing PET QC: QC Image*

Amyloid and tau SUVRs that passed QC timing were additionally QC'd post-processing using a statistical approach, where any scan with an SUVR outside the 1.5 IQR was flagged for visual QC. Additional visual QC was then conducted on a per-cohort basis. If cohort specific amyloid or tau PET values were available (ADNI and A4), we compared our MRI-Free SUVRs with these values and visually inspected any values that deviated from the diagonal. Finally, all tau PET scans that were categorized "Discordant" underwent secondary visual inspection (see below). A total of 47 amyloid scans and 49 tau PET scans were excluded after visual inspection due to normalization issues, cut-offs in the field of view, anatomical abnormalities, or motion (see QC image fails: Figure 1AB).

**Tau PET Positivity Determination**

A gaussian mixture model (GMM) approach was used to ascertain tau positivity (T+/-) across tau PET Braak ROIs using the *mclust* package in R (version 4.4.1) for FTP and MK tau PET scans (e.g. FTP (n=3480 scans from A4, ADNI, Mixed Protocol, and SCAN) and MK (n=1384 scans from WRAP, Mixed Protocol, and SCAN)). This number of tau PET scans represents all available tau scans in the ADSP and is thus higher than the number used in our final analysis. Within each tracer, multiple gaussian distributions were fit to each tau PET ROI (allowing for either equal or unequal variances in these distributions) and the optimal GMM for each ROI was selected by evaluating the Bayesian information criterion (BIC) (Supplemental Figures 4-5). Each participant was assigned a GMM classification of belonging to k=n tau classes for each ROI. The mean and standard deviation (SD) of the lowest tau distribution (i.e. the T- class) was used to reclassify artifactual cases. An example of pre and post reclassification of the Braak 1/2 ROI for FTP scans is provided in Supplemental Figure 6. The threshold for tau positivity for each ROI was determined using the mean and SD of the reclassified T- distribution. Histograms of reclassified distributions from the best fitting GMMs for each ROI for each tracer-specific approach are shown in Supplemental Figures 7-8. A summary of the mean and SD of the reclassified T- distribution and corresponding thresholds of M+2SD and M+2.5SD from the optimal GMM are detailed in Table 3. For all analyses in the manuscript, we used the more conservative M +2.5SD threshold.

We also assessed hierarchical patterns of tau positivity in accordance with Braak staging. For each scan, an algorithm was used to determine each individual's PET-based tau stage in a hierarchical fashion, such that later stages could only be achieved if the individual was tau positive in previous stages (for instance, a T4+ case would also be positive for T12 and T3). Cases that were tau negative across all Braak stage ROIs were labeled T- and any case in which regional positivity did not follow the hierarchical staging schema was labeled “Discordant” (n=169 scans out of 4866 total scans (~3%) in the ADSP) and underwent additional visual QC and review (n=117 included in the current study and detailed in Supplemental Tables 4-5).

**Figure 1. Inclusion and exclusion criteria**

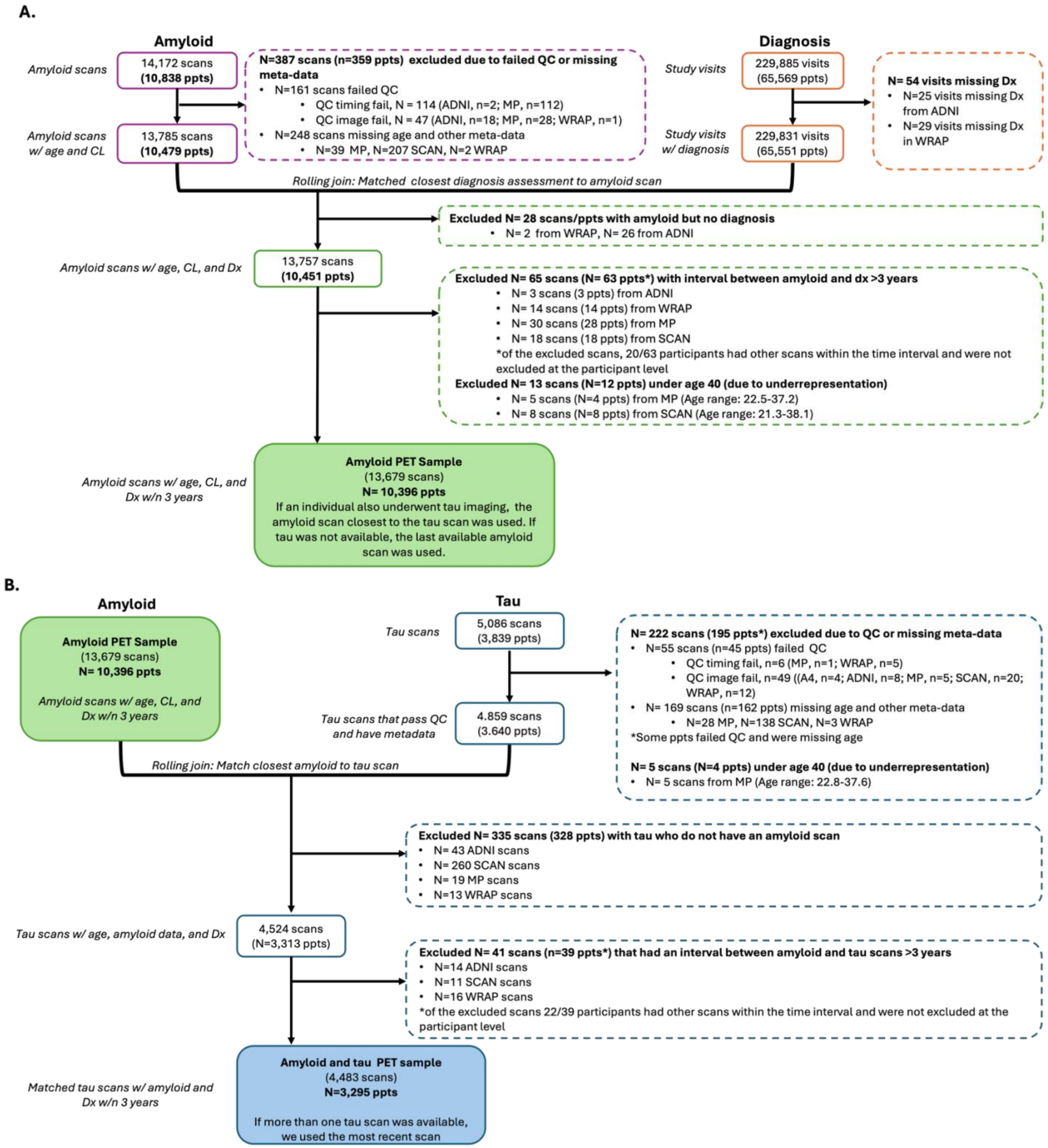


**Figure 1. Inclusion and exclusion criteria for amyloid and tau PET samples.** Panel A shows inclusion criteria for amyloid PET-based frequency analyses. Panel B shows inclusion and exclusion criteria for the tau PET based frequency analyses. Amyloid and tau PET scans were drawn from the ADSP-PHC and SCAN-Prospective datasets. In the amyloid PET sample (A), for individuals with more than one available amyloid scan the amyloid scan closest to the tau scan (if a tau scan existed) was chosen, and if tau was not available for that participant, the most recent amyloid PET scan was used. In the amyloid and tau PET sample (B), for individuals with more than one tau scan, the most recent tau scan was used.

**Figure 2. Tau PET templates**

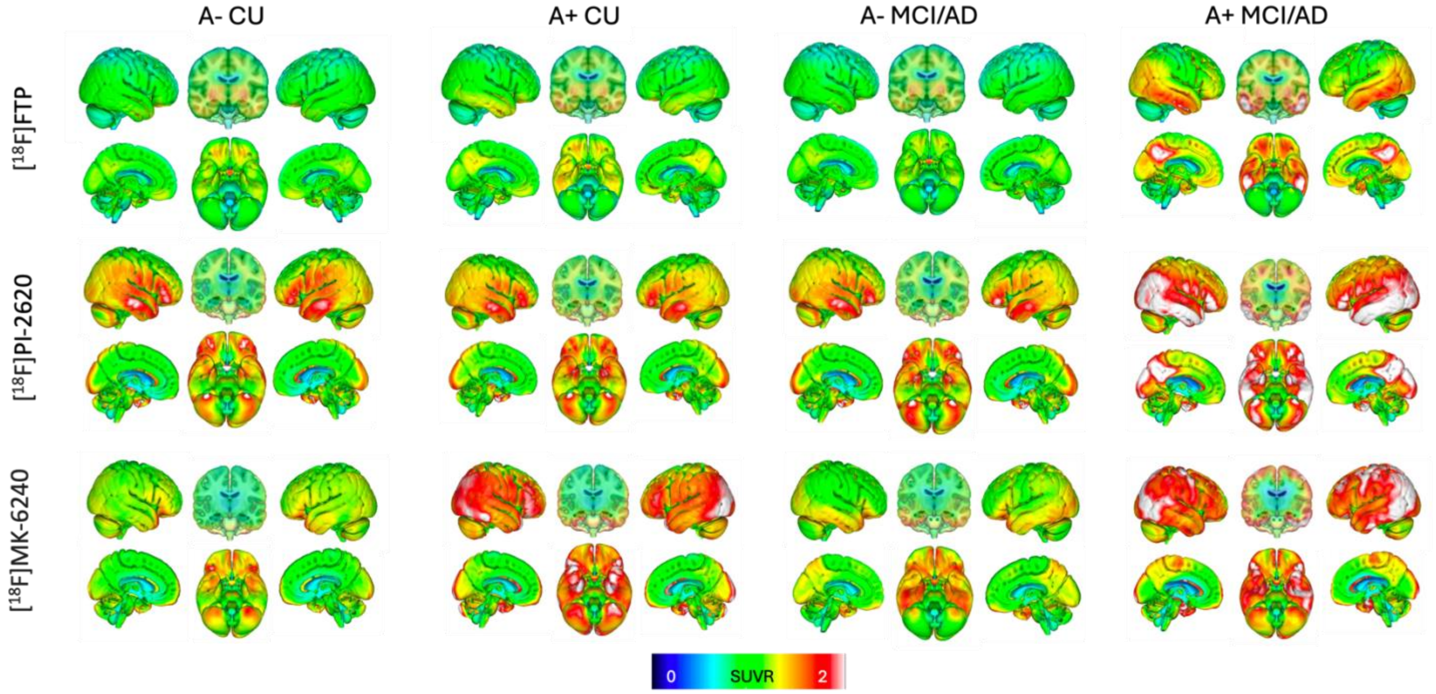


**Figure 2. Tau PET templates.** Surface renderings of the twelve tau PET templates used for non-linear spatial normalization of tau PET data during the MRI-Free processing pipeline are shown above. SUVR templates were generated for four groups based on amyloid status and clinical impairment (e.g., A- CU, A+ CU, A- MCI/AD, and A+ MCI/AD) for each of the three tau PET tracers included in the ADSP-PHC. In the spatial normalization step, all tracer templates were used simultaneously, where SPM linearly combines all the templates for spatial normalization by minimizing a cost function. For the [18F]Flortaucipir templates, 15 subjects were randomly selected per diagnostic group from ADNI data, since A4 did not have any MCI/AD participants. For the Stanford University [18F]PI-2620 templates, 17 subjects were randomly selected out of each of the four diagnostic groups to comprise the templates. This number was chosen because the limiting group was A+ CI, which has a total of 17 subjects. For the [18F]MK-6240 PET templates, 5 subjects' SUVR images were merged for each of the A+ CU and A+ CI templates, and 8 subjects' SUVR images were merged for each the A- CI and A- CU templates. For each template, individual summed PET files were spatially normalized to a T1-weighted 1mm isotropic MNI brain atlas (www.fil.ion.ucl.ac.uk/spm). SUVR images were generated from each normalized PET file by dividing the image by the mean intensity value of the NPDKA-derived inferior cerebellar cortex. SUVR images were summed. The result was smoothed by an 8x8x8 voxel kernel to create the final template image for each tracer specific amyloid and clincial impairment group. CU= Cognitively Unimpaired; MCI = mild cognitive impairment, AD = Alzheimer's dementia; A+/- = amyloid PET positivity.

**Figure 3. Tau PET regions of interest**

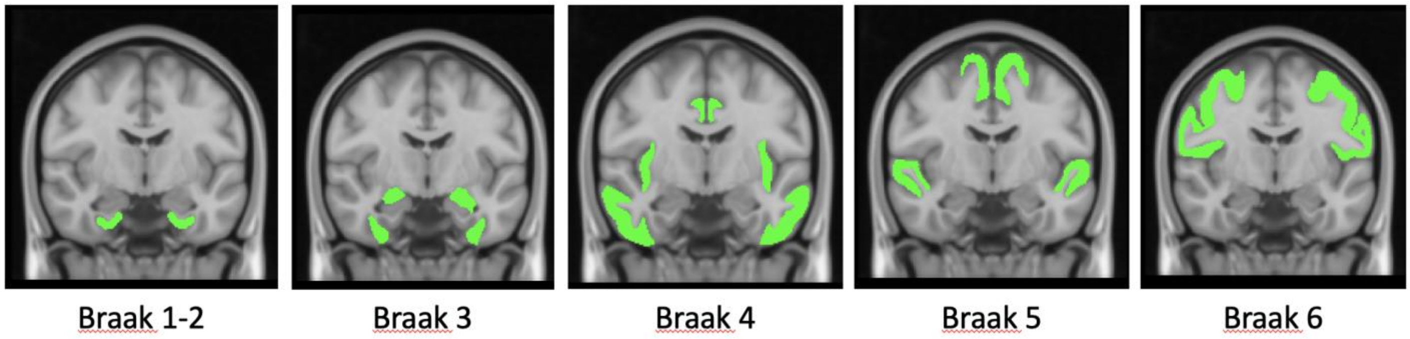


**Figure 3. Tau PET regions of interest.** Tau PET regions corresponding Braak-based neuropathological staging. Braak 1-2: entorhinal cortices; Braak 3: amygdala, parahippocampal gyrus, fusiform gyrus and lingual gyrus; Braak 4: insula, inferior temporal, lateral temporal, posterior cingulate and inferior parietal; Braak 5: superior temporal, precuneus, supramarginal gyrus, superior parietal, frontal (including orbitofrontal, inferior frontal, superior frontal and rostral medial frontal), anterior cingulate, lateral occipital, cuneus; and Braak 6: paracentral, postcentral, precentral and pericalcarine. Hippocampi were excluded from Braak-based ROIs due to high off-target binding in nearby choroid plexus for FTP.

**Figure 4. GMM BIC comparison: FTP**

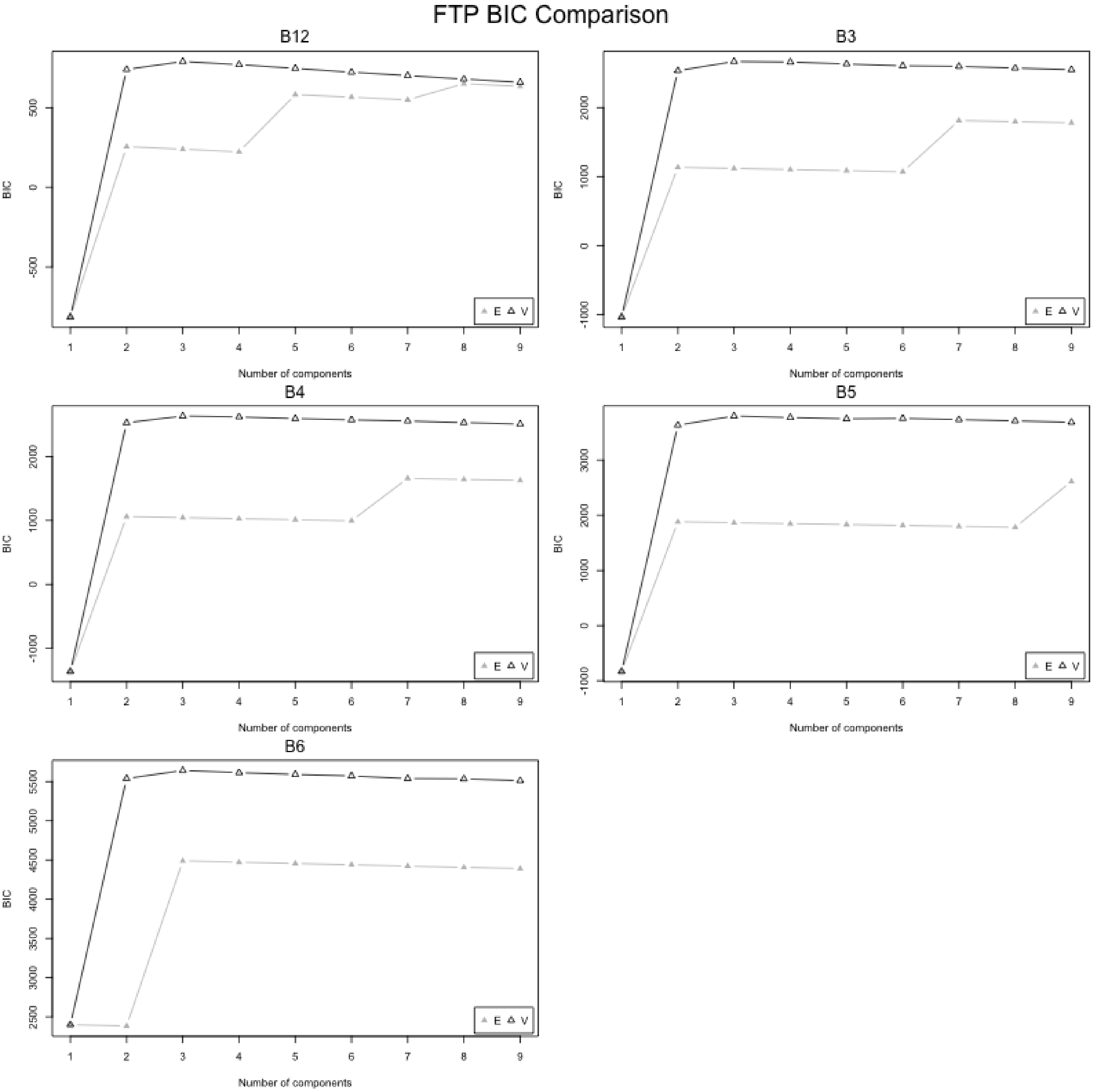


E=Equal variance
V=Unequal variance

**Figure 4. GMM BIC comparison: FTP.** Each plot shows the Bayesian Information Criterion (BIC) values for varying numbers of mixture components (1–9) across FTP-specific Gaussian mixture models (GMMs) for each Braak-based ROI (e.g., B1-B6). Each point represents a fitted GMM with either equal variance (E, gray filled triangles) or unequal variance (V, black empty triangles) across components. Higher BIC values indicate better model fit and clearer class separation, respectively.

**Figure 5. GMM BIC comparison: MK**

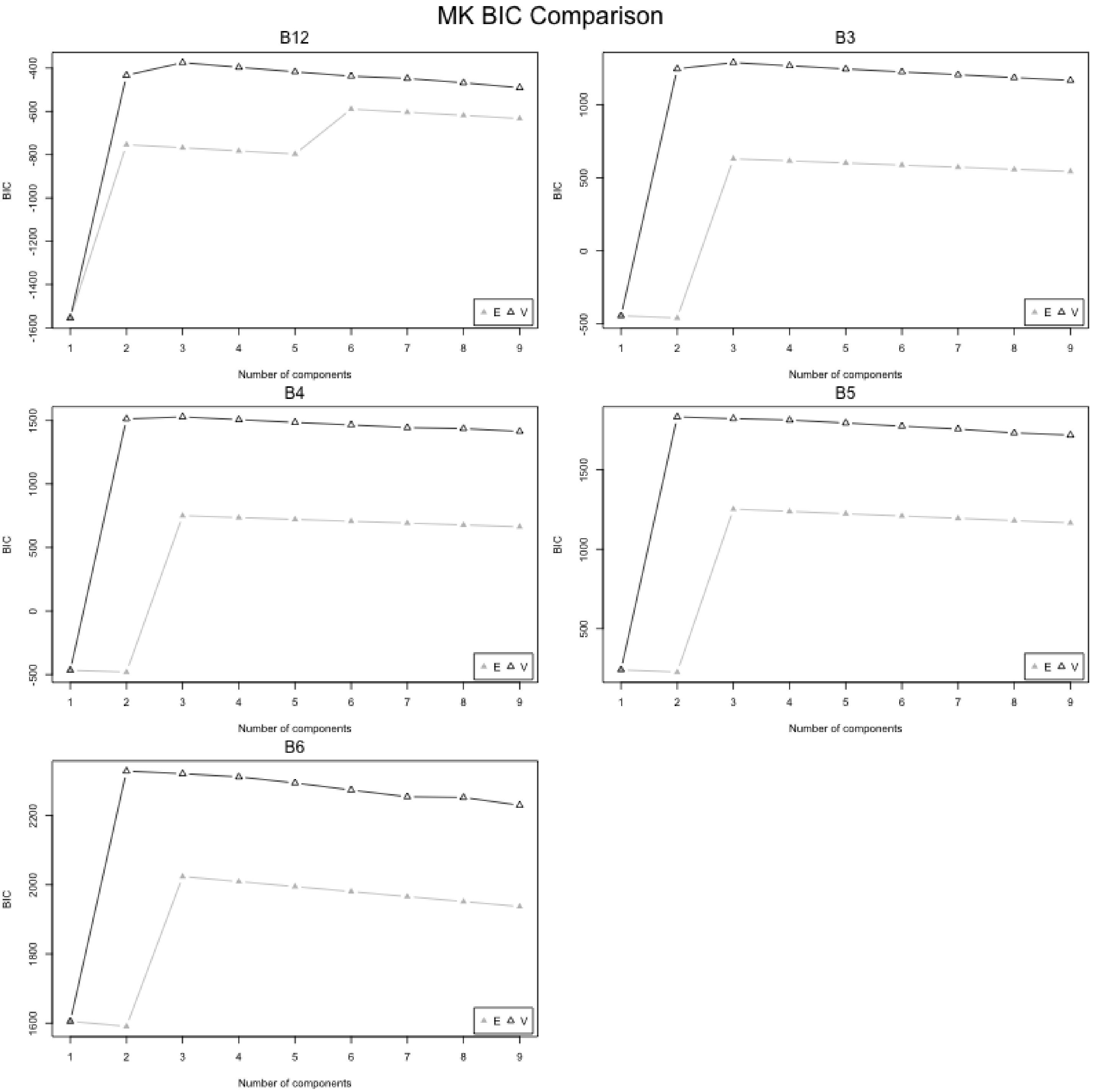


E=Equal variance
V=Unequal variance

**Figure 5. GMM BIC comparison: MK.** Each plot shows the Bayesian Information Criterion (BIC) values for varying numbers of mixture components (1–9) across MK-specific Gaussian mixture models (GMMs) for each Braak-based ROI (e.g., B1-B6). Each point represents a fitted GMM with either equal variance (E, gray filled triangles) or unequal variance (V, black empty triangles) across components. Higher BIC values indicate better model fit and clearer class separation, respectively.

**Figure 6. Example of GMM reclassification**

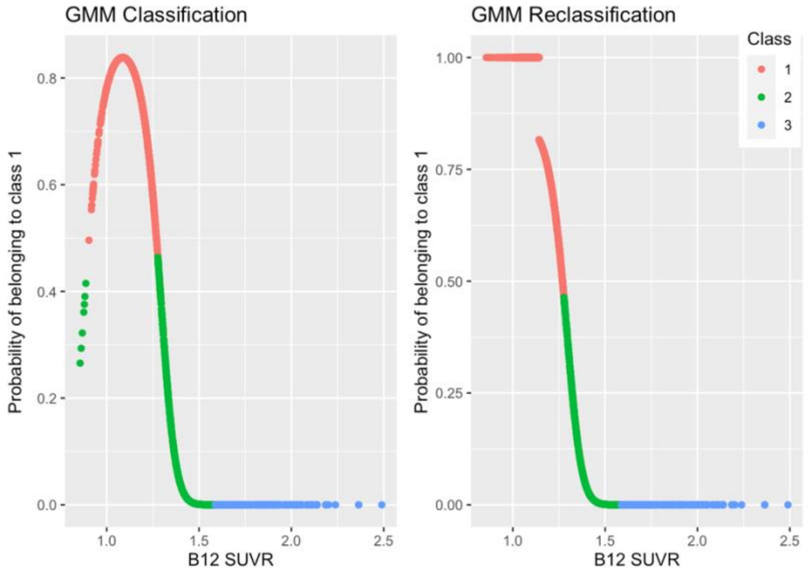


**Figure 6. Example of GMM reclassification.** The results of the optimal GMM for the Braak I/II (B12) ROI for FTP scans included three classes or tau distributions (left panel). The lowest tau distribution (e.g., T- class or class 1) is shown in red and the T+ distributions, classes 2 and 3, are shown in green and blue, respectively. Those belonging to the T+ class(es) with an SUVR value below the mean of the T- class were reclassified as belonging to the T- class, and correspondingly, their probability of belonging to the T- class was set to 100% and probability of belonging to the T+ class was set to 0% (right panel).

**Figure 7. GMM reclassified distributions: FTP scans**

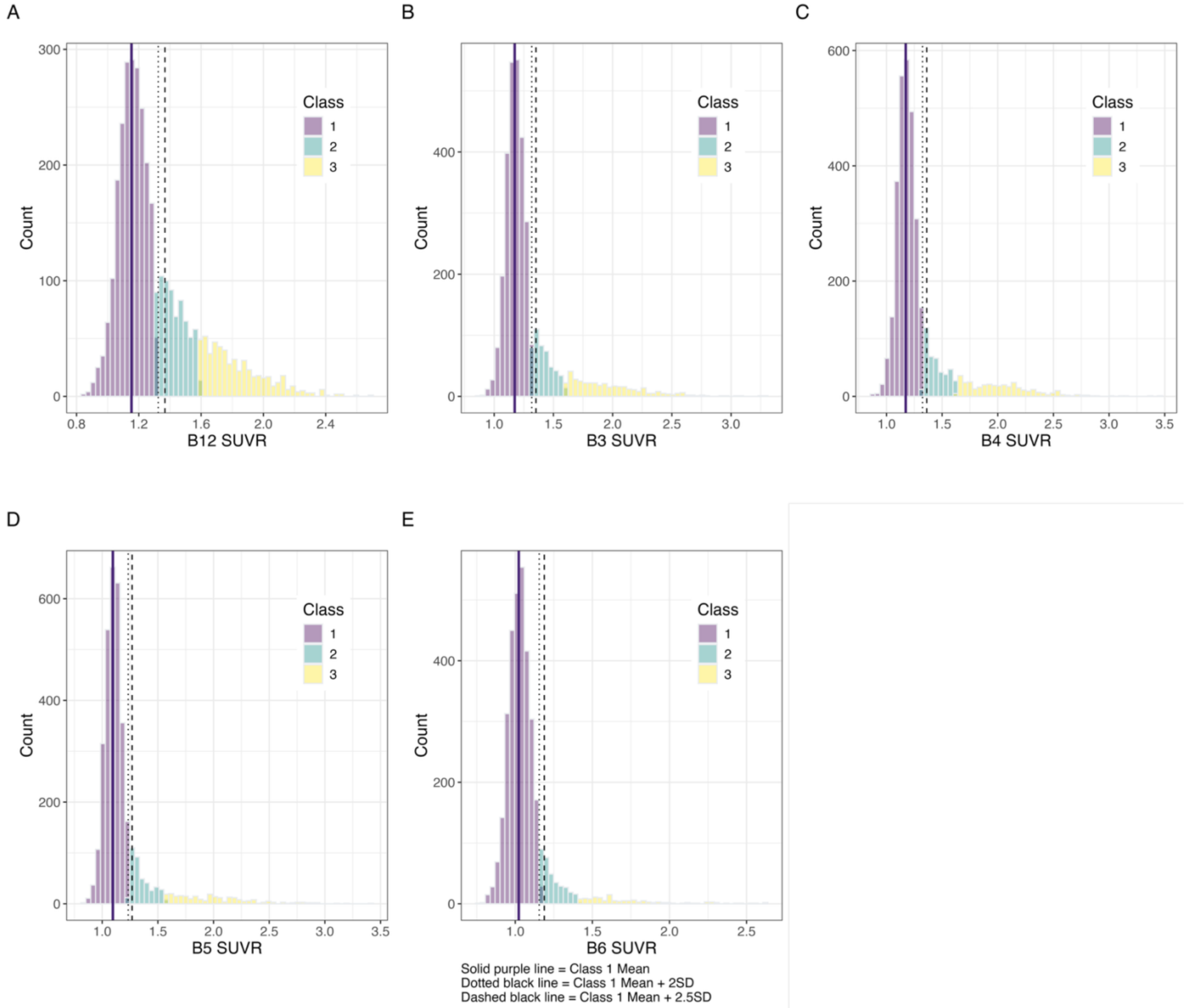


**Figure 7. GMM reclassified distributions: FTP scans.** Histograms of reclassified distributions from the best fitting GMMs for each ROI for the FTP-specific approach (n=3480). The tau negative distribution is shown in purple (e.g. Class 1). The solid purple line indicates the mean, the dotted black line indicates the mean+2SD and the dashed black line indicates the mean+2.5SD of the tau negative distribution for each ROI.

**Figure 8. GMM reclassified distributions: MK scans**

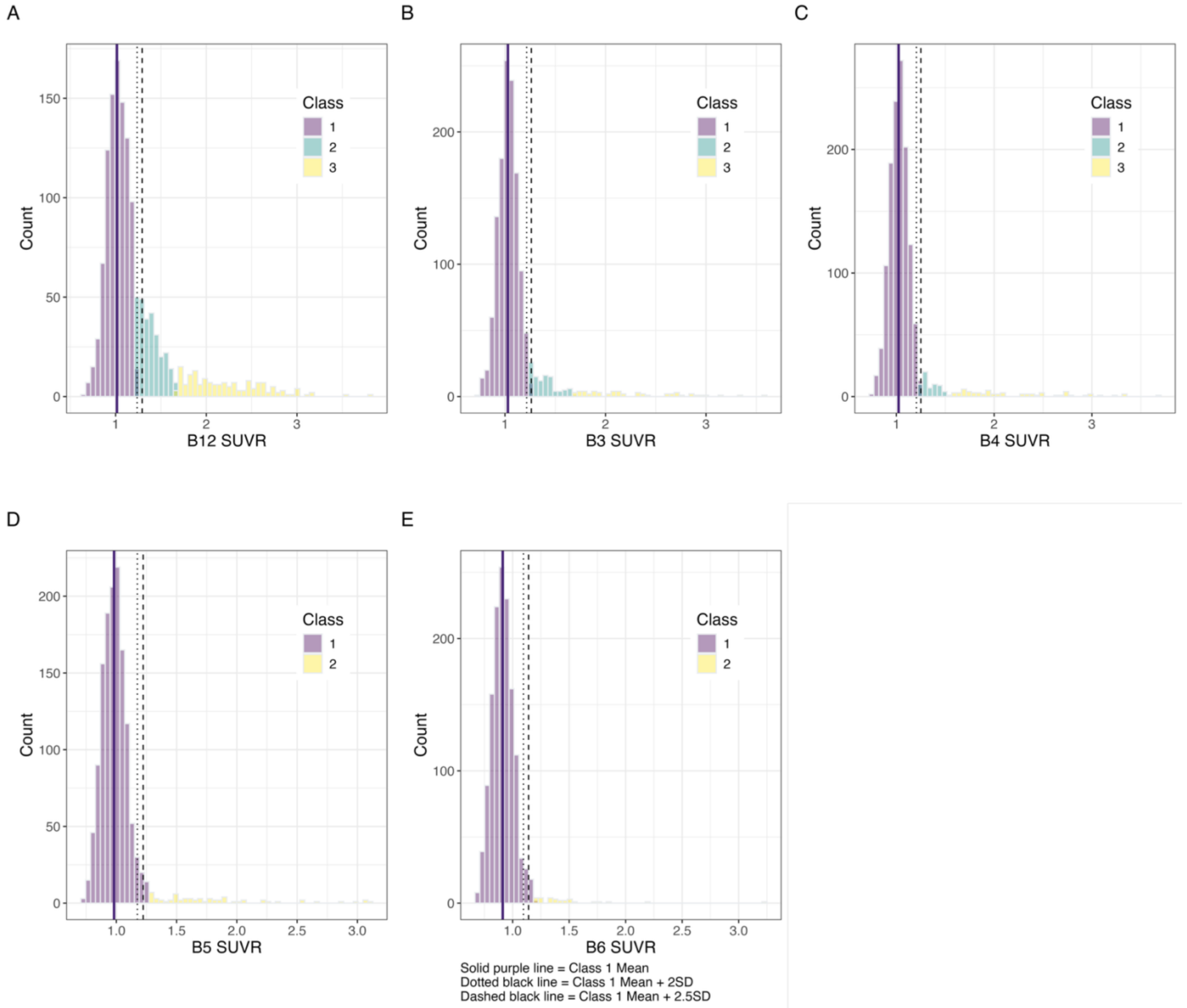


**Figure 8. GMM reclassified distributions: MK scans.** Histograms of reclassified distributions from the best fitting GMMs for each ROI for the MK-specific approach (n=1384). The tau negative distribution is shown in purple (e.g. Class 1). The solid purple line indicates the mean, the dotted black line indicates the mean+2SD and the dashed black line indicates the mean+2.5SD of the tau negative distribution for each ROI.

**Figure 9. Regional tau PET SUVR Z-scores across clinical stages by hierarchical tau PET stage**

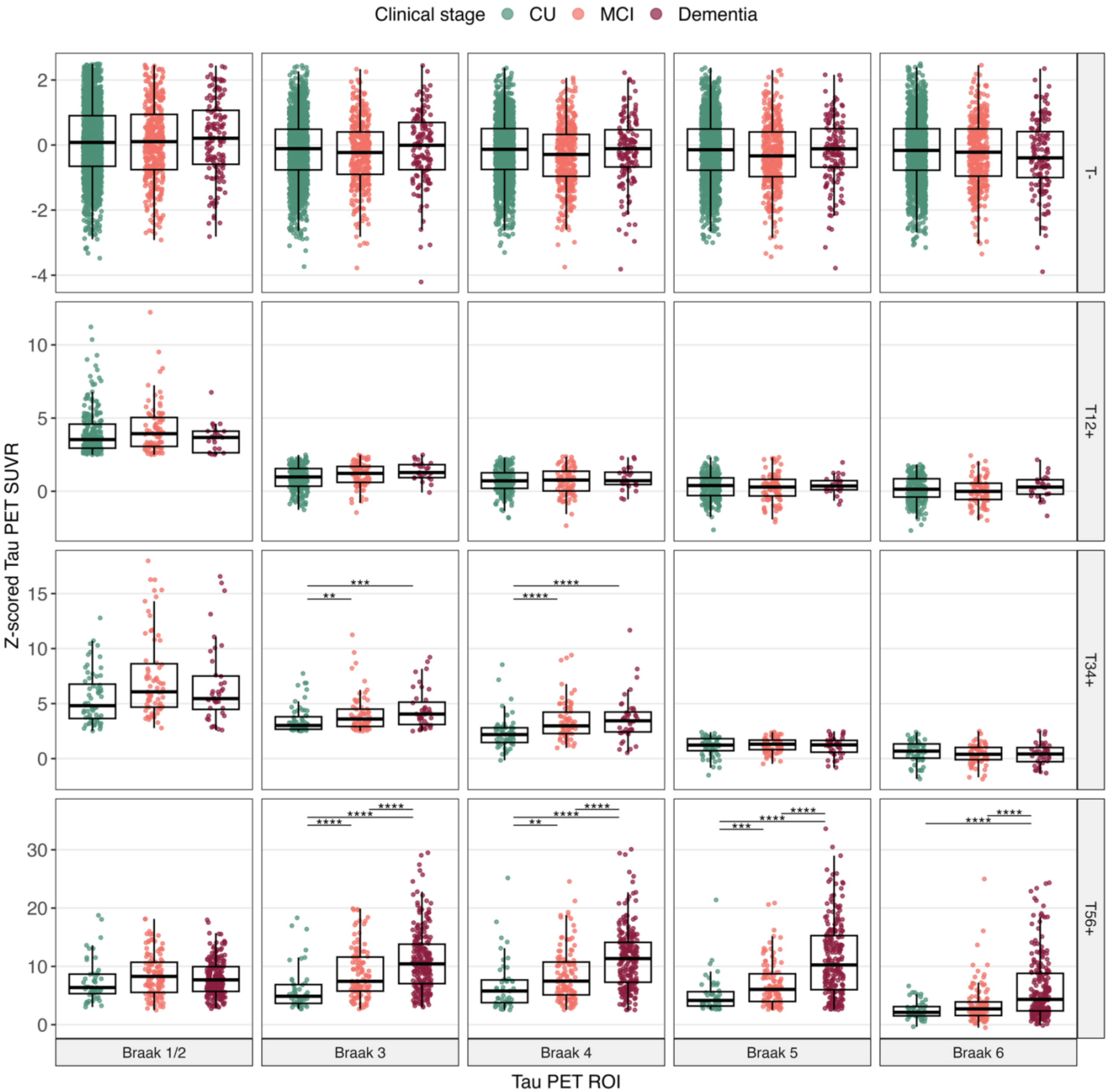


**Figure 9. Regional tau PET SUVR Z-scores across clinical stages by hierarchical tau PET stage.** This figure displays boxplots of Z-scored tau PET SUVR values across Braak-based tau PET ROIs (columns: Braak 1/2 through Braak 6) and stratified by hierarchical tau PET stage (rows: T–, T12+, T34+, T56+). Clinical stages—cognitively unimpaired (CU, green), mild cognitive impairment (MCI, orange), and dementia (maroon)—are shown within each panel. Participant counts (n) per clinical stage for each tau stage are displayed below each panel. Following a significant group test (Kruskal-Wallis), post hoc pairwise comparisons (CU vs MCI, CU vs dementia, MCI vs dementia) were conducted using Mann Whitney U tests with Bonferroni correction. Asterisks denote statistically significant differences after adjustment (*$P$ < .05, **$P$ < .01,***$P$ < .001, ****$P$ < .0001).

| **Table 1. PET timing windows and radiotracers** | | |
|---|---|---|
| Tracer | Compliant Acquisition Start | Compliant Acquisition End |
| FBB | 90 | 110 |
| FBP | 50 | 70 |
| PIB | 50 | 70 |
| FTP* | 80 | 100 |
| MK | 90 | 110 |
| *For ADNI, FTP data corresponding to 75-105 minutes post-injection were processed. Given that the midpoint of the acquisition window is 90 for both 75-105 and 80-100 windows, it is unlikely that the magnitude of SUVR values across these two windows would differ. | | |

**Table 2. Centiloid equations for MRI-Free amyloid PET scans**

| Tracer | MRI-free SUVR → MRI-free CL |
|---|---|
| FBP | (FBP MRI-free CL) = 192.40(MRI-free FBP SUVR) – 207.27 |
| NAV | (NAV MRI-free CL) = 88.86(MRI-free NAV SUVR) – 91.71 |
| FBB | (FBB MRI-free CL) = 164.60(MRI-free FBB SUVR) – 171.97 |
| $PIB_{50\text{-}70}$ | ($PIB_{50to70}$ MRI-free CL) = 92.08(MRI-free PIB SUVR) – 95.83 |
| *Timing adjustment* | |
| $PIB_{50\text{-}70}$[a] | ($PIB_{50to70}$ MRI-free CL) = 1.051(MRI-free $PIB_{40to60}$ CL) + 1.785 |

[a]For the SCAN data, 343 out of 975 PIB scans included in the ADSP were processed using 40-60 min acquisition windows. This linear regression equation was provided by UC Berkeley SCAN PET Core to translate the 40-60min CLs to the corresponding CL value for 50-70 min acquisition window. CL equations and details about the PIB timing window translation are also provided here: https://files.alz.washington.edu/scan/UCBerkeley_SCAN_Amyloid_MRIfree_Methods.pdf

| Table 3. Tracer-specific GMM summary | | | | | | | | |
|---|---|---|---|---|---|---|---|---|
| | | Optimal GMM[a] | | T- class[b] | | | Thresholds | |
| **Tracer** | **ROI** | **Variance** | **No. Classes** | **N** | **Mean** | **SD** | **M+2SD** | **M+2.5SD** |
| **FTP** | B12 | Unequal variance | 3 | 2200 | 1.153 | 0.086 | 1.325 | 1.368 |
| **(ADNI + A4 + MP + SCAN)** | B3 | Unequal variance | 3 | 2600 | 1.173 | 0.072 | 1.316 | 1.352 |
| N=3480 scans | B4 | Unequal variance | 3 | 2697 | 1.173 | 0.076 | 1.324 | 1.362 |
| | B5 | Unequal variance | 3 | 2818 | 1.096 | 0.069 | 1.267 | 1.268 |
| | B6 | Unequal variance | 3 | 3005 | 1.023 | 0.066 | 1.155 | 1.188 |
| **MK** | B12 | Unequal variance | 3 | 954 | 1.014 | 0.111 | 1.237 | 1.293 |
| **(WRAP + MP+ SCAN)** | B3 | Unequal variance | 3 | 1226 | 1.029 | 0.093 | 1.215 | 1.262 |
| N=1384 scans | B4 | Unequal variance | 3 | 1261 | 1.023 | 0.091 | 1.205 | 1.250 |
| | B5 | Unequal variance | 2 | 1322 | 0.982 | 0.097 | 1.175 | 1.223 |
| | B6 | Unequal variance | 2 | 1356 | 0.912 | 0.092 | 1.095 | 1.141 |

[a]Optimal GMM for each ROI was selected using the Bayesian Information Criterion (Figures 4-5).
[b]The mean and SD reported here are rounded to the thousandths place; thus, the corresponding thresholds, which were calculated without rounding, may not add up exactly when estimated from the table M and SD values due to rounding.

| Table 4. Summary of quantitatively discordant tau PET cases (n = 117) | | | |
|---|---|---|---|
| **Visual tau pattern category** | **Reason for disagreement between visual and quantitative assessment (if any)** | **Quantitative tau positivity pattern** | **Visual tau positivity pattern** |
| **A. Visually concordant tau PET patterns (n=68)** | | | |
| **Visually tau negative** (n=42) | Threshold (n=23) | 3 (n=7), 356 (n=1), 36 (n=1), 4 (n=4), 5 (n=3), 6 (n=8) | 0 (n=23) |
| | Off target (n=18) | 1235 (n=1), 12356 (n=1), 1236 (n=1), 124 (n=1), 34 (n=1), 356 (n=1), 4 (n=1), 456 (n=4), 5 (n=3), 56 (n=1), 6 (n=3) | 0 (n=18) |
| | Technical issue: Uptake in reference region and off target (n=1) | 56 (n=1) | 0 (n=1) |
| **Visually hierarchical pattern** (n=26) | Threshold (n=9) | 12356 (n=1), 124 (n=3), 126 (n=1), 3 (n=1), 345 (n=1), 3456 (n=1) | 1234 (n=5), 12345 (n=2), 123456 (n=2) |
| | Asymmetry (n=9) | 12356 (n=1), 124 (n=2), 1245 (n=3), 34 (n=3) | 1234 (n=5), 12345 (n=3), 123456 (n=1) |
| | Off target (n=6) | 12356 (n=1), 124 (n=1), 1245 (n=2), 126 (n=2) | 12 (n=5), 1234 (n=1) |
| | Technical issue: Reference region uptake (n=1), misaligned ROI (n=1) | 124 (n=1), 3 (n=1) | 1234 (n=1), 123 (n=1) |
| **B. Visually discordant tau PET patterns (n=49)** | | | |
| **MTL sparing** (Sparing Braak 12 or 123; n=34) | Agreement between quantitative and visual pattern (n=19) | 3 (n=1), 34 (n=2), 345 (n=2), 3456 (n=6), 5 (n=3), 56 (n=5) | 3 (n=1), 34 (n=2), 345 (n=2), 3456 (n=6), 5 (n=3), 56 (n=5) |
| | Atrophy (signal diluted due to atrophy; n=4) | 345 (n=1), 56 (n=1), 356 (n=1), 3456 (n=1) | 3456 (n=2), 34 (n=1), 45 (n=1) |
| | Asymmetry (n=11) | 3 (n=2), 4 (n=5), 356 (n=1), 456 (n=1), 5 (n=1), 12356 (n=1) | 34 (n=6), 345 (n=1), 3456 (n=1), 45 (n=1), 56 (n=2) |
| **Atypical pattern** (n=15) | Agreement between quantitative and visual pattern (n=11) | 1235 (n=1), 12356 (n=2), 124 (n=5), 1245 (n=1), 125 (n=2) | 1235 (n=1), 12356 (n=2), 124 (n=5), 1245 (n=1), 125 (n=2) |
| | Asymmetry and/or focal uptake (n=4) | 12456 (n=1), 4 (n=1), 5 (n=1), 1256 (n=1) | 12346 (n=1), 14 (n=1), 15 (n=1), 15 (n=1) |
| A summary of visual tau PET patterns and comparison to quantitative tau PET patterns for a total of 117 cases that had non-hierarchical or "discordant" tau. Cases were categorized into visually concordant (A: Visually tau negative or visually hierarchical) and visually discordant (B: MTL sparing and atypical patterns). Reasons for differences in visual and quantitative tau patterns as well as the quantitative tau PET pattern (e.g. 123, corresponding to T+ in Braak 1-2 and Braak 3 ROIs and 0, corresponds to T- across all ROIs) and the visually observed tau PET pattern are provided in each column. | | | |

**Table 5. Characteristics of participants with quantitatively discordant tau patterns**

| | | Discordant tau PET category | | | |
|---|---|---|---|---|---|
| | | Visually concordant | | Visually discordant | |
| **Characteristic** | **Overall** N = 117 | **Tau negative** N = 42 | **Hierarchical** N = 26 | **MTL-sparing** N = 34 | **Atypical** N = 15 |
| **Tau tracer** | | | | | |
| FTP | 97 (83%) | 25 (60%) | 24 (92%) | 34 (100%) | 14 (93%) |
| MK | 20 (17%) | 17 (40%) | 2 (8%) | 0 (0%) | 1 (7%) |
| **Imaging cohort** | | | | | |
| A4 | 16 (14%) | 3 (7%) | 4 (15%) | 8 (24%) | 1 (7%) |
| ADNI | 36 (31%) | 8 (19%) | 7 (27%) | 14 (41%) | 7 (47%) |
| MP | 9 (8%) | 1 (2%) | 4 (15%) | 4 (12%) | 0 (0%) |
| SCAN | 40 (34%) | 17 (40%) | 9 (35%) | 8 (24%) | 6 (40%) |
| WRAP | 16 (14%) | 13 (31%) | 2 (8%) | 0 (0%) | 1 (7%) |
| **Age at tau PET** | 72.7 (7.1) | 71.5 (6.6) | 75.3 (7.2) | 74.3 (6.3) | 68.2 (7.9) |
| **Amyloid PET positivity** | 72 (62%) | 8 (19%) | 24 (92%) | 25 (74%) | 15 (100%) |
| **Diagnosis** | | | | | |
| CU | 69 (59%) | 35 (83%) | 11 (42%) | 15 (44%) | 8 (53%) |
| MCI | 34 (29%) | 1 (2%) | 6 (23%) | 7 (21%) | 0 (0%) |
| Dementia | 14 (12%) | 6 (14%) | 9 (35%) | 12 (35%) | 7 (47%) |
| **Female** | 62 (53%) | 25 (60%) | 13 (50%) | 11 (32%) | 13 (87%) |
| **APOE4 carriers** | 36 (47%) | 5 (24%) | 9 (45%) | 13 (50%) | 9 (90%) |

Values shown as n (%) or Mean (SD).

Abbreviations: A4, Anti-Amyloid Treatment in Asymptomatic Alzheimer disease; ADNI, Alzheimer's disease Neuroimaging Initiative; APOE, apolipoprotein; CU, Cognitively unimpaired; FTP, Flortaucipir; MCI, mild cognitive impairment; MK, MK6240; SCAN, Standardized Centralized Alzheimer's & Related Dementias Neuroimaging; WRAP, Wisconsin Registry for Alzheimer's Prevention

**Table 6. Amyloid PET imaging sample participant characteristics by imaging cohort**

| Characteristic | Overall N = 10,396 | A4 N = 4,486 | ADNI N = 1,806 | NACC[a] Mixed Protocol N=1,504 | NACC[a] SCAN N=2,010 | WRAP N = 590 |
|---|---|---|---|---|---|---|
| **Tracer** | | | | | | |
| FBB | 1,306 (13%) | 0 (0%) | 385 (21%) | 152 (10%) | 769 (38%) | 0 (0%) |
| FBP | 6,738 (65%) | 4,486 (100%) | 1,357 (75%) | 639 (42%) | 256 (13%) | 0 (0%) |
| NAV | 133 (1.3%) | 0 (0%) | 0 (0%) | 0 (0%) | 107 (5.3%) | 26 (4.4%) |
| PIB | 2,219 (21%) | 0 (0%) | 64 (3.5%) | 713 (47%) | 878 (44%) | 564 (96%) |
| **Age at amyloid scan (years)** | 71.89 (7.05) | 71.29 (4.68) | 75.39 (8.07) | 70.53 (8.55) | 72.08 (7.97) | 68.59 (7.24) |
| **Clinical stage** | | | | | | |
| CU | 7,764 (75%) | 4,486 (100%) | 708 (39%) | 832 (55%) | 1,201 (60%) | 537 (91%) |
| MCI | 1,480 (14%) | 0 (0%) | 674 (37%) | 296 (20%) | 465 (23%) | 45 (7.6%) |
| Dementia | 1,152 (11%) | 0 (0%) | 424 (23%) | 376 (25%) | 344 (17%) | 8 (1.4%) |
| **Female** | 5,948 (57%) | 2,663 (59%) | 881 (49%) | 846 (56%) | 1,158 (58%) | 400 (68%) |
| ***APOE*4 Carriage**[b] | | | | | | |
| e4- | 5,424 (62%) | 2,891 (65%) | 923 (57%) | 688 (61%) | 669 (61%) | 253 (62%) |
| e4+ | 3,269 (38%) | 1,550 (35%) | 705 (43%) | 435 (39%) | 427 (39%) | 152 (38%) |
| **Education (years)**[c] | 16.44 (2.74) | 16.58 (2.84) | 16.26 (2.59) | 16.34 (2.87) | 16.38 (2.50) | 16.43 (2.82) |
| **Race**[d] | | | | | | |
| American Indian or Alaska Native | 59 (0.6%) | 9 (0.2%) | 3 (0.2%) | 11 (0.7%) | 36 (1.8%) | 0 (0%) |
| Asian | 299 (2.9%) | 170 (3.8%) | 46 (2.5%) | 30 (2.0%) | 52 (2.6%) | 1 (0.2%) |
| Black | 613 (5.9%) | 159 (3.5%) | 2 (0.1%) | 77 (5.1%) | 333 (17%) | 42 (7.2%) |
| Multiple, Other, Unknown | 143 (1.4%) | 53 (1.2%) | 37 (2.0%) | 25 (1.7%) | 26 (1.3%) | 2 (0.3%) |
| Native Hawaiian or Other Pacific Islander | 139 (1.3%) | 2 (<0.1%) | 134 (7.4%) | 1 (<0.1%) | 2 (0.1%) | 0 (0%) |
| White | 9,126 (88%) | 4,093 (91%) | 1,584 (88%) | 1,360 (90%) | 1,551 (78%) | 538 (92%) |
| **Ethnicity**[e] | | | | | | |
| Hispanic or Latino | 463 (4.5%) | 142 (3.2%) | 98 (5.4%) | 94 (6.3%) | 127 (6.3%) | 2 (0.3%) |
| Not Hispanic or Latino | 9,880 (96%) | 4,309 (97%) | 1,701 (95%) | 1,410 (94%) | 1,881 (94%) | 579 (100%) |

Values shown as n (%) or Mean (SD).

[a] NACC includes PET images from Mixed Protocol and SCAN (Supplemental Table 4)

[b] A total of 8,693 had available genetic data (n, missing = 1,703)

[c] A total of 10,357 had available education data (n, missing = 39)

[d] A total of 10,379 had available race data (n, missing = 17)

[e] A total of 10,343 had available ethnicity data (n, missing = 53)

Abbreviations: A4, Anti-Amyloid Treatment in Asymptomatic Alzheimer disease; ADNI, Alzheimer's disease Neuroimaging Initiative; APOE, apolipoprotein; CU, Cognitively unimpaired; FBB, 18F-Florbetaben; FBP, 18F-Florbetapir; MCI, mild cognitive impairment; NACC, National Alzheimer's Coordinating Center; NAV, 18F-NAV-4694; PIB, 11C-Pittsburgh Compound B; SCAN, Standardized Centralized Alzheimer's & Related Dementias Neuroimaging; WRAP, Wisconsin Registry for Alzheimer's Prevention

**Table 7. Observed probabilities of amyloid positivity**

| | Clinical stage | | |
|---|---|---|---|
| **Age group** | **CU** | **MCI** | **Dementia** |
| [42.5,47.5) | 16.7 (1/6) | - | - |
| [47.5,52.5) | 3.7 (1/27) | 27.3 (3/11) | 60.0 (9/15) |
| [52.5,57.5) | 6.7 (6/90) | 24.2 (8/33) | 78.3 (47/60) |
| [57.5,62.5) | 9.7 (23/236) | 31.8 (27/85) | 69.5 (91/131) |
| [62.5,67.5) | 22.4 (371/1658) | 40.3 (79/196) | 61.6 (90/146) |
| [67.5,72.5) | 26.4 (717/2715) | 49.4 (157/318) | 74.9 (140/187) |
| [72.5,77.5) | 31.0 (538/1737) | 52.8 (190/360) | 81.0 (200/247) |
| [77.5,82.5) | 37.4 (348/931) | 63.2 (158/250) | 78.1 (164/210) |
| [82.5,87.5) | 35.9 (99/276) | 51.3 (78/152) | 83.5 (81/97) |
| [87.5,92.5) | 37.9 (25/66) | 55.9 (33/59) | 70.2 (33/47) |
| [92.5,97.5) | 21.1 (4/19) | 41.7 (5/12) | 100.0 (7/7) |
| [97.5, 102.5] | - | - | - |

Data are observed probabilities in % (No. amyloid positive/No. total subgroup). No estimates were provided if the age group included <5 participants.

**Table 8. Predicted probabilities of amyloid positivity**

| | Clinical stage | | |
|---|---|---|---|
| **Age group** | **CU** | **MCI** | **Dementia** |
| 45 | 8.0 (6.3–9.9) | - | - |
| 50 | 10.0 (8.3–12.0) | 32.2 (25.5–39.6) | 66.5 (58.6–73.5) |
| 55 | 12.6 (10.9–14.5) | 36.2 (30.3–42.5) | 69.2 (63.1–74.7) |
| 60 | 15.7 (14.1–17.4) | 40.4 (35.4–45.6) | 71.8 (67.2–76.0) |
| 65 | 19.3 (17.9–20.9) | 44.8 (40.8–48.8) | 74.3 (70.8–77.4) |
| 70 | 23.6 (22.3–25.0) | 49.2 (46.0–52.5) | 76.6 (73.8–79.2) |
| 75 | 28.5 (27.1–30.1) | 53.7 (50.7–56.7) | 78.7 (75.9–81.3) |
| 80 | 34.0 (31.9–36.2) | 58.1 (54.6–61.6) | 80.8 (77.5–83.6) |
| 85 | 39.9 (36.9–43.0) | 62.4 (57.9–66.7) | 82.6 (78.8–85.9) |
| 90 | 46.2 (42.2–50.3) | 66.5 (61.0–71.6) | 84.3 (79.8–88.0) |
| 95 | 52.6 (47.5–57.6) | 70.4 (63.8–76.2) | 85.9 (80.8–89.9) |

Predicted probabilities and 95% CI for A+ are shown as percentages. The probability estimates were generated from a logistic regression model that included age ($P$<.001), clinical stage ($P$<.001), cohort ($P$<.001), and age by clinical stage ($P$<.001) as predictors. No estimates were provided if the age group included <5 participants. The median difference between the predicted and observed probabilities of A+ was 4.6% [IQR: 0.1% to 9.0%] across age bin comparisons indicating good model fit (Tables 6-7).

| **Table 9. Odds ratios from binomial logistic regression predicting amyloid positivity** | | | |
|---|---|---|---|
| | **OR** | **95% CI** | **P value** |
| **Clinical stage (Reference=CU)** | | | |
| MCI | 3.14 | 2.71–3.63 | <.001 |
| Dementia | 10.56 | 8.97–12.46 | <.001 |
| **Age** | 1.05 | 1.04–1.06 | <.001 |
| **Cohort (Reference=A4)** | | | |
| ADNI | 0.74 | 0.64–0.86 | <.001 |
| NACC | 0.65 | 0.57–0.73 | <.001 |
| WRAP | 0.74 | 0.60–0.91 | .005 |
| **Age x Clinical stage** | | | |
| Age x MCI | 0.98 | 0.97-1.00 | .05 |
| Age x Dementia | 0.97 | 0.96-0.99 | .002 |

The odds ratios (ORs) were generated from a binomial logistic regression model of amyloid positivity (versus amyloid negativity) and included age (mean-centered), clinical stage, cohort, and age by clinical stage as predictors. ORs greater than 1 indicate increased odds of amyloid positivity; values less than 1 indicate decreased odds.

**Table 10. Logistic model comparisons for amyloid outcomes**

**A. Models compared predicting amyloid positivity (Logistic regression models)**

| Model terms | AIC | LRT | df | P value |
|---|---|---|---|---|
| Age + clinical stage + cohort | 12261 | 10.7 | 2 | 0.005 |
| Age × clinical stage + cohort | 12255 | | | |

**B. Models compared predicting amyloid levels (Ordinal logistic regression models)**

| Model terms | AIC | LRT | df | P value |
|---|---|---|---|---|
| Age + clinical stage + cohort | 29346 | 15.3 | 2 | <0.001 |
| Age × clinical stage + cohort | 29335 | | | |

Abbreviations: AIC, Akaike information criterion; LRT, log-likelihood ratio test; df, degrees of freedom
[a]Models predicting amyloid outcomes were compared using AIC and log-likelihood ratio tests (LRT; $c^2$) with p-values degrees of freedom (df) reported. All models contained lower levels terms. Age was mean-centered. Positive LRT indicates that the interactive model had lower deviance and better model fit than the additive model.

**Table 11. Odds ratios from ordinal logistic regression predicting amyloid levels**

| | OR | 95% CI | P value |
|---|---|---|---|
| **Clinical stage (Reference=CU)** | | | |
| MCI | 3.08 | 2.70-3.52 | <.001 |
| Dementia | 12.17 | 10.57-14.01 | <.001 |
| **Age** | 1.04 | 1.03-1.05 | <.001 |
| **Cohort (Reference=A4)** | | | |
| ADNI | 0.66 | 0.58-0.75 | <.001 |
| NACC | 0.62 | 0.57-0.69 | <.001 |
| WRAP | 0.65 | 0.55-0.77 | <.001 |
| **Age x Clinical stage** | | | |
| Age x MCI | 1.00 | 0.99-1.01 | .95 |
| Age x Dementia | 0.97 | 0.96-0.99 | <.001 |

The odds ratios (ORs) were generated from an ordinal logistic regression model estimating the odds of being in a higher amyloid level (e.g., 6 CL-based amyloid levels) and included age (mean-centered), clinical stage, cohort, and age by clinical stage as predictors. The ORs indicate the change in odds of being in a higher amyloid stage per unit change in the predictor. OR > 1 = higher odds of more severe amyloid levels. OR < 1 = lower odds of more severe amyloid levels.

**Table 12. Tau PET imaging sample participant characteristics by cohort**

| Characteristic | Overall N = 3,295 | A4 N = 443 | ADNI N = 925 | NACC-ADRC Mixed Protocol N = 352 | NACC-ADRC SCAN N = 1,044 | WRAP N = 531 |
|---|---|---|---|---|---|---|
| **Amyloid Tracer** | | | | | | |
| FBB | 563 (17%) | 0 (0%) | 361 (39%) | 18 (5.1%) | 184 (18%) | 0 (0%) |
| FBP | 1,085 (33%) | 443 (100%) | 564 (61%) | 31 (8.8%) | 47 (4.5%) | 0 (0%) |
| NAV | 123 (3.7%) | 0 (0%) | 0 (0%) | 0 (0%) | 102 (9.8%) | 21 (4.0%) |
| PIB | 1,524 (46%) | 0 (0%) | 0 (0%) | 303 (86%) | 711 (68%) | 510 (96%) |
| **Tau Tracer** | | | | | | |
| FTP | 2,347 (71%) | 443 (100%) | 925 (100%) | 294 (84%) | 685 (66%) | 0 (0%) |
| MK | 948 (29%) | 0 (0%) | 0 (0%) | 58 (16%) | 359 (34%) | 531 (100%) |
| **Age at tau scan (years)** | 71.88 (8.07) | 71.82 (4.84) | 74.91 (8.12) | 68.14 (9.66) | 71.41 (8.04) | 70.08 (7.19) |
| **Time between amyloid and tau scans (years)** | 0.29 (0.61) | 0.00 (0.00) | 0.23 (0.60) | 0.06 (0.31) | 0.11 (0.45) | 1.15 (0.57) |
| **Clinical stage** | | | | | | |
| CU | 2,156 (65%) | 443 (100%) | 498 (54%) | 112 (32%) | 620 (59%) | 483 (91%) |
| MCI | 697 (21%) | 0 (0%) | 310 (34%) | 95 (27%) | 250 (24%) | 42 (7.9%) |
| Dementia | 442 (13%) | 0 (0%) | 117 (13%) | 145 (41%) | 174 (17%) | 6 (1.1%) |
| **Female** | 1,893 (57%) | 254 (57%) | 482 (52%) | 188 (53%) | 612 (59%) | 357 (67%) |
| ***APOE*4 Status**[a] | | | | | | |
| E4- | 1,382 (58%) | 204 (47%) | 469 (61%) | 182 (60%) | 309 (59%) | 218 (61%) |
| E4+ | 1,009 (42%) | 232 (53%) | 303 (39%) | 120 (40%) | 213 (41%) | 141 (39%) |
| **Education (years)** | 16.35 (2.64) | 16.21 (2.84) | 16.41 (2.46) | 16.48 (2.95) | 16.26 (2.49) | 16.48 (2.85) |
| **Amyloid Positive** | 1,404 (43%) | 349 (79%) | 381 (41%) | 153 (43%) | 387 (37%) | 134 (25%) |

Values shown as n (%) or Mean (SD)

[a]N=2,391 had available *APOE4* data (n=904 missing)

[b]N=3,276 had available education data (n=19 missing)

Abbreviations: A4, Anti-Amyloid Treatment in Asymptomatic Alzheimer disease; ADNI, Alzheimer's disease Neuroimaging Initiative; APOE, apolipoprotein; CU, Cognitively unimpaired; FBB, 18F-Florbetaben; FBP, 18F-Florbetapir; FTP, 18F-Flortaucipir; MCI, mild cognitive impairment; MK, 18F-MK-6240; NACC, National Alzheimer's Coordinating Center; NAV, 18F-NAV-4694; PIB, 11C-Pittsburgh Compound B; SCAN, Standardized Centralized Alzheimer's & Related Dementias Neuroimaging; WRAP, Wisconsin Registry for Alzheimer's Prevention

**Table 13. Logistic model comparisons for tau stage outcomes**

**Models compared predicting tau stage (Ordinal logistic regression models)**

| **Model terms** | **AIC** | **LRT** | **df** | **P value** |
|---|---|---|---|---|
| **a.** Centiloids + Age + clinical stage + cohort | 4071.6 | a vs b: 55.2 | 2 | <.001 |
| **b**. Centiloids + Age × clinical stage + cohort | 4020.4 | | | |
| **c**. Centiloids × clinical stage + Age × clinical stage + cohort | 3968.0 | b vs c: 56.4 | 2 | <.001 |
| **d.** Centiloids × clinical stage + Age × clinical stage + centiloids × clinical stage cohort | 3843.0 | c vs d: 126.5 | 1 | <.001 |
| **e.** Age × centiloids × clinical stage + cohort | 3834.9 | d vs e: 12.6 | 2 | .002 |

Abbreviations: AIC, Akaike information criterion; LRT, log-likelihood ratio test; df, degrees of freedom

[a]Models predicting the probability of tau stages were compared using AIC and log-likelihood ratio tests (LRT; $c^2$) with p-values degrees of freedom (df) reported. All models contained lower levels terms. Age and centiloids were mean centered. Positive LRT indicates that the second model (b in a vs b) had lower deviance and a better model fit.

| **Table 14. Odds ratios from ordinal logistic regression predicting tau PET stages** | | | |
|---|---|---|---|
| | **OR** | **95% CI** | ***P* value** |
| **Clinical stage (Reference=CU)** | | | |
| MCI | 2.424 | 1.746-3.327 | <.001 |
| Dementia | 6.583 | 4.519-9.495 | <.001 |
| **Age** | 1.064 | 1.045-1.083 | <.001 |
| **Centiloids** | 1.032 | 1.028-1.037 | <.001 |
| **Cohort (Reference=A4)** | | | |
| ADNI | 2.488 | 1.700-3.676 | <.001 |
| NACC | 3.299 | 2.282-4.828 | <.001 |
| WRAP | 4.448 | 2.993-6.676 | <.001 |
| **Age x Clinical stage** | | | |
| Age x MCI | 0.999 | 0.956-1.019 | <.001 |
| Age x Dementia | 0.915 | 0.881-0.951 | <.001 |
| **Age x Centiloids** | 0.999 | 0.998-0.999 | <.001 |
| **Centiloids x Clinical stage** | | | |
| Centiloids x MCI | 1.018 | 1.011-1.026 | 0.41 |
| Centiloids x Dementia | 1.025 | 1.017-1.034 | <.001 |
| **Age x Centiloids x Clinical stage** | | | |
| Age x Centiloids x MCI | 0.999 | 0.998-0.999 | <.001 |
| Age x Centiloids x Dementia | 0.999 | 0.999-1.000 | 0.22 |

The odds ratios (ORs) were generated from an ordinal logistic regression model estimating the odds of being in a higher tau stage (e.g., T-, T12+, T34+, T56+) and included age (mean-centered), centiloids (centered on 25), clinical stage, all their 2- and 3-way interactions, as well as cohort as predictors. The ORs indicate the change in odds of being in a higher tau stage per unit change in the predictor. OR > 1 = higher odds of more severe tau stages. OR < 1 = lower odds of more severe tau stages.

| **Table 15. Estimated centiloids at which the T12 and T34 probability curves peak across clinical stages[a]** | | | | |
|---|---|---|---|---|
| **A. Comparison of peak centiloids between T12 and T34 within clinical stages** | | | | |
| | **T12+ Peak Centiloid** Mean (95% CI) | **T34+ Peak Centiloid** Mean (95% CI) | **T34 vs T12 Peak Centiloid** Mean difference (95% CI) | **P value** |
| **CU** | 100 (89-113) | 137 (121-157) | 37 (32-43) | <.001 |
| **MCI** | 54 (49-60) | 78 (72-85) | 24 (21-27) | <.001 |
| **Dementia** | 34 (28-39) | 54 (48-60) | 20 (17-23) | <.001 |

| **B. Pairwise comparisons of peak centiloids across clinical stages within each tau stage** | | | |
|---|---|---|---|
| **Tau stage** | **Clinical stage comparison** | **Mean difference in peak centiloids (95% CI)** | **P value** |
| **T12** | CU - MCI | 45 (33-60) | <.001 |
| | CU - Dementia | 66 (54-80) | <.001 |
| | MCI - Dementia | 21 (13-29) | <.001 |
| **T34** | CU - MCI | 58 (43-79) | <.001 |
| | CU - Dementia | 83 (66-103) | <.001 |
| | MCI - Dementia | 24 (17-34) | <.001 |

[a]Peak centiloids refer to the centiloid value at which the predicted probability for a given tau stage is maximized, based on an ordinal logistic regression model which included centiloids (mean-centered), age (mean-centered), clinical stage and their 2- and 3-way interactions as well as cohort, all of which were statistically significant ($P < .001$; see Table 12). Marginal predicted probabilities were estimated using the ggeffects package in R. For each tau stage and clinical stage, the centiloid value at which the predicted probability peaked was extracted. To assess uncertainty and perform group comparisons, we implemented a bootstrap procedure with 1,000 resamples, in which the model was re-fit on each resample. For each comparison, we computed the mean difference in peak centiloids, 95% bootstrap confidence intervals (CIs), and two-sided bootstrap *P* values.

**Table 16. Logistic model comparisons for biological stage outcomes**

**Models compared predicting amyloid levels (Ordinal logistic regression models)**

| **Model terms** | **AIC** | **LRT** | **df** | **P value** |
|---|---|---|---|---|
| Age + clinical stage + cohort | 6540.4 | 69.9 | 2 | <.001 |
| Age × clinical stage + cohort | 6474.4 | | | |

Abbreviations: AIC, Akaike information criterion; LRT, log-likelihood ratio test; df, degrees of freedom
[a]Models predicting prevalence of biological stages were compared using AIC and log-likelihood ratio tests (LRT; $c^2$) with p-values degrees of freedom (df) reported. All models contained lower levels terms. Age was mean centered. Positive LRT indicates that the synergistic model had lower deviance and better model fit than the additive model.

**Table 17. Odds ratios from ordinal logistic regression predicting biological stages**

| | OR | 95% CI | P value |
|---|---|---|---|
| **Clinical stage (Reference=CU)** | | | |
| MCI | 3.97 | 3.18 -4.95 | <.001 |
| Dementia | 30.74 | 23.57-40.26 | <.001 |
| **Age** | 1.07 | 1.06-1.08 | <.001 |
| **Cohort (Reference=A4)** | | | |
| ADNI | 0.20 | 0.15-0.25 | <.001 |
| NACC | 0.21 | 0.17-0.26 | <.001 |
| WRAP | 0.25 | 0.19-0.32 | <.001 |
| **Age x Clinical stage** | | | |
| Age x MCI | 0.98 | 0.96-1.01 | 0.158 |
| Age x Dementia | 0.90 | 0.88-0.93 | <.001 |

The odds ratios (ORs) were generated from an ordinal logistic regression model estimating the odds of being in a higher biological stage and included age (mean-centered), clinical stage, cohort, and age by clinical stage as predictors. The ORs indicate the change in odds of being in a higher biological stage per unit change in the predictor. OR > 1 = higher odds of more severe biological stages. OR < 1 = lower odds of more severe biological stages.
